# Equity and Transportability of Plasma ATN Phenotypes in a Population-Representative U.S. Aging Cohort

**DOI:** 10.64898/2026.01.31.26344775

**Authors:** Emmanuel Fle Chea

**Affiliations:** Independent Researcher, University of Minnesota School of Public Health (Alumnus), Minneapolis, MN, USA

**Keywords:** Alzheimer’s disease, plasma biomarkers, ATN framework, health equity, transportability, fairness metrics, structural disadvantage, vascular comorbidity

## Abstract

**INTRODUCTION:** Plasma biomarkers for Alzheimer’s disease (AD) pathology promise scalable diagnostic access, yet their performance in diverse, population-representative cohorts remains uncharacterized. We evaluated equity and transportability of plasma amyloid-tau-neurodegeneration (ATN) biomarkers in a nationally representative U.S. aging cohort.

**METHODS:** Cross-sectional analysis of 4,427 adults aged ≥50 years from the 2016 Health and Retirement Study Venous Blood Study. Plasma biomarkers (Aβ42/40, pTau181, NfL, GFAP) were classified using established ATN criteria. Survey weights produced population-representative estimates. Outcomes included biomarker-cognition associations, fairness metrics (sensitivity, specificity, predictive values) stratified by race/ethnicity and sex, and education-stratified analyses.

**RESULTS:** Among 4,427 participants representing 36.6 million U.S. adults (weighted: 68 years, 55% female, 79% White), survey-weighted analysis revealed tau as the only biomarker maintaining robust cognitive associations (β=**-**0.74, p<0.001), while amyloid (β=0.11, p=0.43) and neurodegeneration (β=**-**0.27, p=0.08) lost significance. White participants demonstrated 12-percentage-point higher sensitivity than Black participants (23.4% vs. 11.4%), with Black women showing lowest sensitivity (8.8%). Educational attainment modified biomarker effects: low-education groups showed paradoxical positive amyloid associations (β=0.74, p=0.01) and amplified neurodegeneration effects (β=**-**1.02, p=0.006). Race-specific optimal cutpoints differed by 40%. Vascular comorbidity burden was higher in Black (82%) and Hispanic (73%) versus White (65%) participants, yet associations persisted after vascular adjustment.

**DISCUSSION:** Plasma ATN biomarkers demonstrate significant equity gaps and differential transportability across demographic subgroups. The 12-percentage-point sensitivity disparity and education-dependent effect modification highlight barriers to equitable implementation. Population-based validation with fairness metrics should be prerequisite for clinical deployment.

## INTRODUCTION

Plasma biomarkers for Alzheimer’s disease (AD) pathology, including amyloid-β 42/40 ratio, phosphorylated tau 181 (pTau181), neurofilament light (NfL), and glial fibrillary acidic protein (GFAP), enable scalable implementation of the amyloid-tau-neurodegeneration (ATN) framework previously restricted to cerebrospinal fluid and positron emission tomography studies [1**-**3]. These blood-based assays promise to democratize access to AD pathology assessment, particularly for underrepresented populations historically excluded from specialized research centers [4].

However, most plasma biomarker validation studies derive from highly selected clinical cohorts characterized by European ancestry, high educational attainment, and low comorbidity burden [5-7]. These convenience samples introduce selection bias that threatens transportability, the extent to which research findings generalize to target populations [11]. Without population-representative data, plasma biomarker thresholds and clinical utility metrics may systematically misrepresent diverse communities, perpetuating rather than mitigating health disparities [16,17].

Three critical gaps limit current understanding. First, the transportability of plasma ATN phenotypes from research cohorts to general populations remains uncharacterized. Survey-weighted analysis can reveal whether unweighted estimates typical in published literature accurately reflect population-level burden [12]. Second, fairness of biomarker performance across demographic subgroups requires quantification using machine learning equity metrics (true positive rate, false positive rate, positive/negative predictive value) [16,18,19]. Third, structural disadvantage, operationalized through educational attainment as a proxy for life-course socioeconomic adversity, may modify biomarker-cognition relationships [21,22].

The Health and Retirement Study (HRS), a nationally representative longitudinal cohort of U.S. adults aged ≥50 years with plasma biomarker data from 4,427 participants, provides an unprecedented opportunity to address these gaps [8,9]. Unlike clinic-based cohorts, HRS employs probability sampling with survey weights enabling population-level inference [10].

We evaluated plasma ATN biomarkers in HRS with three objectives: (1) assess transportability by comparing weighted versus unweighted prevalence and associations; (2) quantify fairness disparities across race/ethnicity, sex, and intersections; and (3) examine education as a marker of structural disadvantage modifying biomarker-cognition relationships. We hypothesized that survey-weighted estimates would reveal distinct population-level patterns, fairness metrics would expose differential performance across demographic groups, and educational stratification would illuminate structural mechanisms underlying disparities.

## METHODS

### Study Population

The Health and Retirement Study is a nationally representative longitudinal panel of U.S. adults aged ≥50 years employing multistage probability sampling with oversampling of Black and Hispanic individuals [8]. We analyzed data from the 2016 HRS Venous Blood Study, which collected plasma samples from 9,934 participants [9]. After excluding those with missing biomarker data (n=38), our analytic sample comprised 4,427 individuals with complete ATN classification and cognitive assessment.

### Plasma Biomarker Measurement and ATN Classification

Plasma biomarkers (Aβ42/40 ratio, pTau181, NfL, GFAP) were measured using Simoa assays following standardized protocols [9]. ATN classification followed established criteria [3]: A (Amyloid): Aβ42/40 < 0.063; T (Tau): pTau181 > 2.5 pg/mL; N (Neurodegeneration): NfL > 20 pg/mL OR GFAP > 100 pg/mL. Participants were classified into eight ATN profiles (A**-**T**-**N**-**, A+T**-**N**-**, A**-**T+N**-**, A**-**T**-**N+, A+T+N**-**, A+T**-**N+, A**-**T+N+, A+T+N+).

### Cognitive Assessment

Cognitive function was assessed using the Telephone Interview for Cognitive Status modified (TICSm), a 27-point scale [25]. Cognitive impairment was classified as: dementia (TICSm ≤6), cognitive impairment no dementia (CIND; TICSm 7**-**11), or normal cognition (TICSm ≥12) [26].

### Demographic and Socioeconomic Variables

Race/ethnicity was categorized as non-Hispanic White, non-Hispanic Black, Hispanic, or Other. Educational attainment was measured in years and categorized as: less than high school (<12 years), high school graduate (12 years), some college (13**-**15 years), or college graduate (≥16 years). For interaction models, we created a binary indicator for low education (<12 years) representing structural disadvantage [21]. Intersectional groups were created by crossing race/ethnicity with sex. Vascular comorbidities (hypertension, diabetes, stroke) were self-reported physician diagnoses.

### Survey Weighting

HRS employs complex multistage probability sampling with stratification and clustering. We utilized person-level survey weights (PVBSWGTR) adjusting for sampling probability, non-response, and post-stratification [10]. All descriptive statistics and regression models used survey-weighted methods via the R survey package [13] to produce population-representative estimates. Unweighted estimates were computed for transportability comparison.

### Statistical Analysis

#### Transportability Framework

We compared weighted versus unweighted estimates for: (1) ATN prevalence across profiles; (2) biomarker-cognition associations from multivariable linear regression models adjusting for age, sex, race/ethnicity, and education. Differences between weighted and unweighted estimates quantify selection bias [11,12].

#### Fairness Metrics

We defined “predicted high risk” as A+T+ (AD pathology) and “actual outcome” as cognitive impairment (CIND or dementia). Standard classification metrics were calculated by race/ethnicity, sex, and intersectional subgroups: true positive rate (TPR/sensitivity), false positive rate (FPR), positive predictive value (PPV), and negative predictive value (NPV) [16,19]. Disparities were quantified as absolute differences in TPR between demographic groups.

#### Education and Structural Disadvantage

We conducted: (1) education-stratified ATN prevalence; (2) education-stratified regression models; (3) interaction models testing biomarker × education interactions on cognitive performance [21,22].

#### Race × Biomarker Interactions

We fitted regression models with race/ethnicity × biomarker interaction terms to assess differential effects across groups.

#### Vascular Comorbidity Adjustment

We fitted models with and without adjustment for vascular comorbidities to assess whether cerebrovascular burden confounds biomarker-cognition associations. We also stratified by cardiovascular disease (CVD) presence.

#### Sensitivity Analyses

We conducted: (1) varying ATN cutpoints to evaluate robustness; (2) inverse probability weighting for missingness; (3) bootstrap confidence intervals (1,000 iterations) [14]; (4) Youden-optimized cutpoints by race/ethnicity; (5) biomarker distribution comparisons. Details in Supplementary Methods.

All analyses used R version 4.3.2. Two-sided p-values <0.05 were considered statistically significant. No corrections for multiple comparisons were applied given exploratory nature of equity analyses [15].

## RESULTS

### Sample Characteristics

Among 4,427 participants with complete data, the unweighted sample comprised 59.2% women, 16.7% Black, 14.9% Hispanic, and 65.2% White individuals (Table 1). Mean age was 68.3 years. After applying survey weights representing 36.6 million U.S. adults aged ≥50 years, the weighted sample comprised 54.6% women, 8.8% Black, 9.0% Hispanic, and 78.9% White, reflecting HRS oversampling design. Biomarker missingness was minimal (<1% overall), with pTau181 showing highest missingness (0.79%).

**Table 1.**
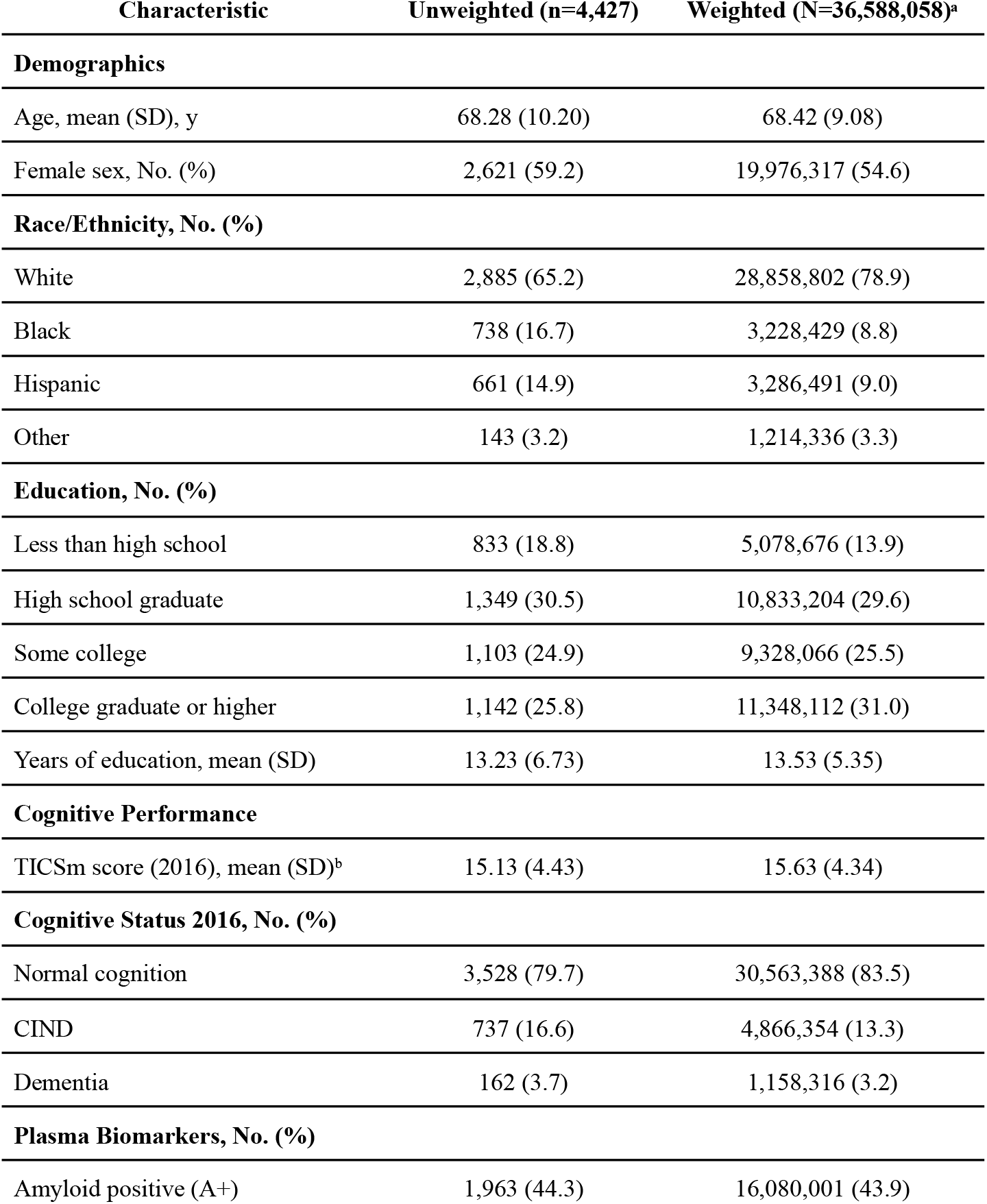
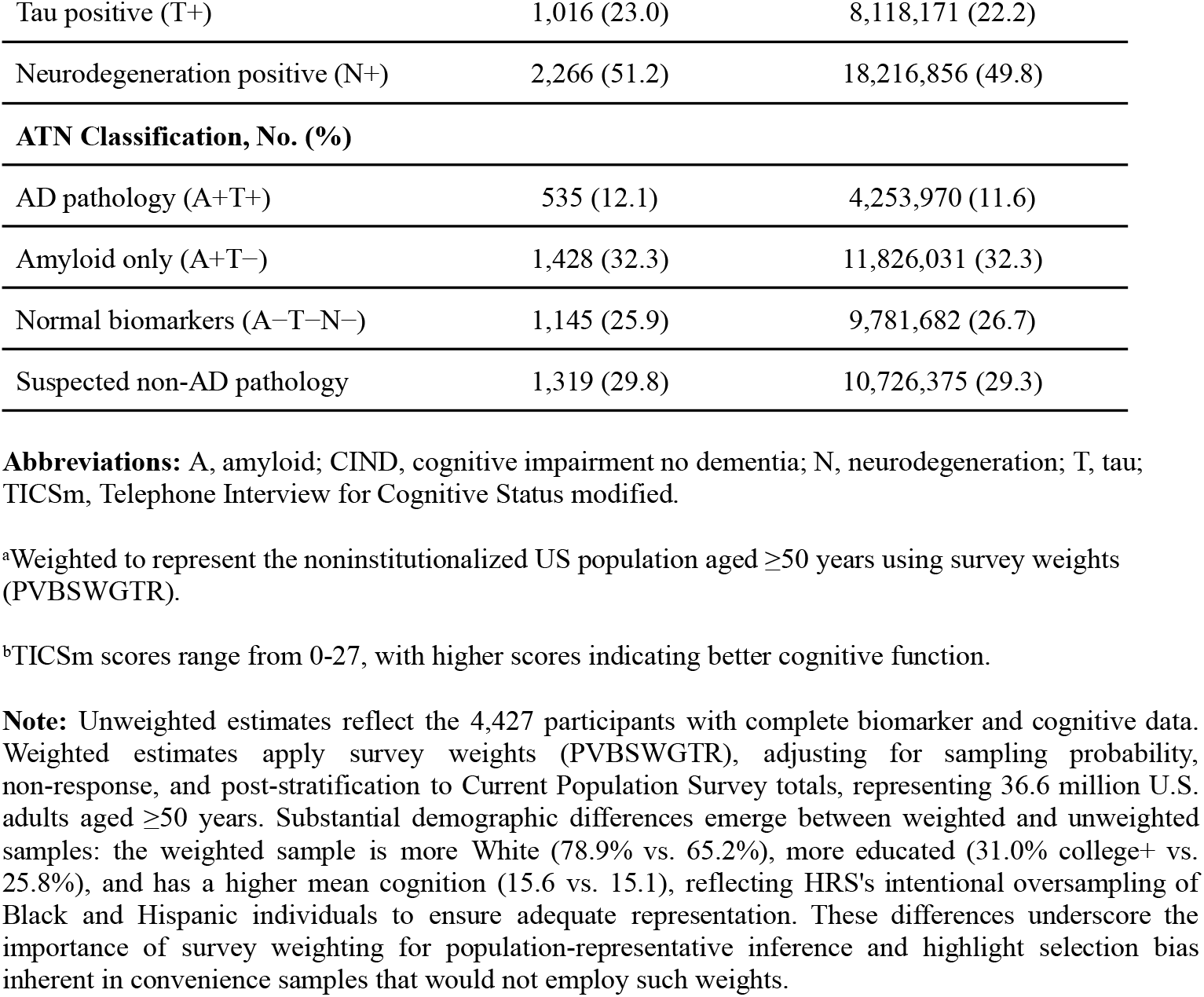
Sample Characteristics: Unweighted and Weighted Estimates from the 2016 Health and Retirement Study Venous Blood Study.

| Characteristic | Unweighted (n=4,427) | Weighted (N=36,588,058)<sup>a</sup> |
| --- | --- | --- |
| **Demographics** |  |  |
| Age, mean (SD), y | 68.28 (10.20) | 68.42 (9.08) |
| Female sex, No. (%) | 2,621 (59.2) | 19,976,317 (54.6) |
| **Race/Ethnicity, No. (%)** |  |  |
| White | 2,885 (65.2) | 28,858,802 (78.9) |
| Black | 738 (16.7) | 3,228,429 (8.8) |
| Hispanic | 661 (14.9) | 3,286,491 (9.0) |
| Other | 143 (3.2) | 1,214,336 (3.3) |
| **Education, No. (%)** |  |  |
| Less than high school | 833 (18.8) | 5,078,676 (13.9) |
| High school graduate | 1,349 (30.5) | 10,833,204 (29.6) |
| Some college | 1,103 (24.9) | 9,328,066 (25.5) |
| College graduate or higher | 1,142 (25.8) | 11,348,112 (31.0) |
| Years of education, mean (SD) | 13.23 (6.73) | 13.53 (5.35) |
| **Cognitive Performance** |  |  |
| TICSm score (2016), mean (SD)<sup>b</sup> | 15.13 (4.43) | 15.63 (4.34) |
| **Cognitive Status 2016, No. (%)** |  |  |
| Normal cognition | 3,528 (79.7) | 30,563,388 (83.5) |
| CIND | 737 (16.6) | 4,866,354 (13.3) |
| Dementia | 162 (3.7) | 1,158,316 (3.2) |
| **Plasma Biomarkers, No. (%)** |  |  |
| Amyloid positive (A+) | 1,963 (44.3) | 16,080,001 (43.9) |
| Tau positive (T+) | 1,016 (23.0) | 8,118,171 (22.2) |
| Neurodegeneration positive (N+) | 2,266 (51.2) | 18,216,856 (49.8) |
| **ATN Classification, No. (%)** |  |  |
| AD pathology (A+T+) | 535 (12.1) | 4,253,970 (11.6) |
| Amyloid only (A+T-) | 1,428 (32.3) | 11,826,031 (32.3) |
| Normal biomarkers (A-T-N-) | 1,145 (25.9) | 9,781,682 (26.7) |
| Suspected non-AD pathology | 1,319 (29.8) | 10,726,375 (29.3) |

**Abbreviations:** A, amyloid; CIND, cognitive impairment no dementia; N, neurodegeneration; T, tau; TICS<sub>m</sub>, Telephone Interview for Cognitive Status modified.
<sup>a</sup>Weighted to represent the noninstitutionalized US population aged $\geq 50$ years using survey weights (PVBSWGTR).
<sup>b</sup>TICS<sub>m</sub> scores range from 0-27, with higher scores indicating better cognitive function.
**Note:** Unweighted estimates reflect the 4,427 participants with complete biomarker and cognitive data. Weighted estimates apply survey weights (PVBSWGTR), adjusting for sampling probability, non-response, and post-stratification to Current Population Survey totals, representing 36.6 million U.S. adults aged $\geq 50$ years. Substantial demographic differences emerge between weighted and unweighted samples: the weighted sample is more White (78.9% vs. 65.2%), more educated (31.0% college+ vs. 25.8%), and has a higher mean cognition (15.6 vs. 15.1), reflecting HRS's intentional oversampling of Black and Hispanic individuals to ensure adequate representation. These differences underscore the importance of survey weighting for population-representative inference and highlight selection bias inherent in convenience samples that would not employ such weights.

### Transportability: Weighted vs. Unweighted Biomarker-Cognition Associations

Biomarker-cognition associations demonstrated substantial transportability differences (Table 2; Figure 3). In unweighted models, tau (β=**-**0.82, p<0.001) and neurodegeneration (β=**-**0.49, p<0.001) showed significant negative associations with cognition, while amyloid was borderline (β=0.21, p=0.06).

**Table 2.**
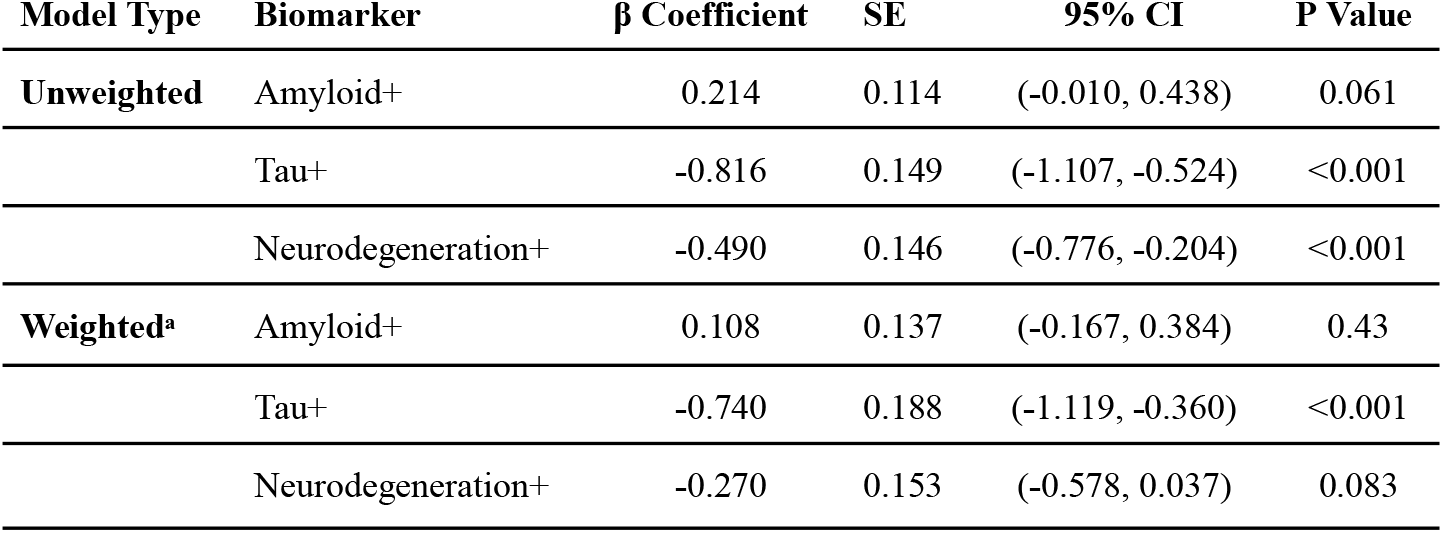
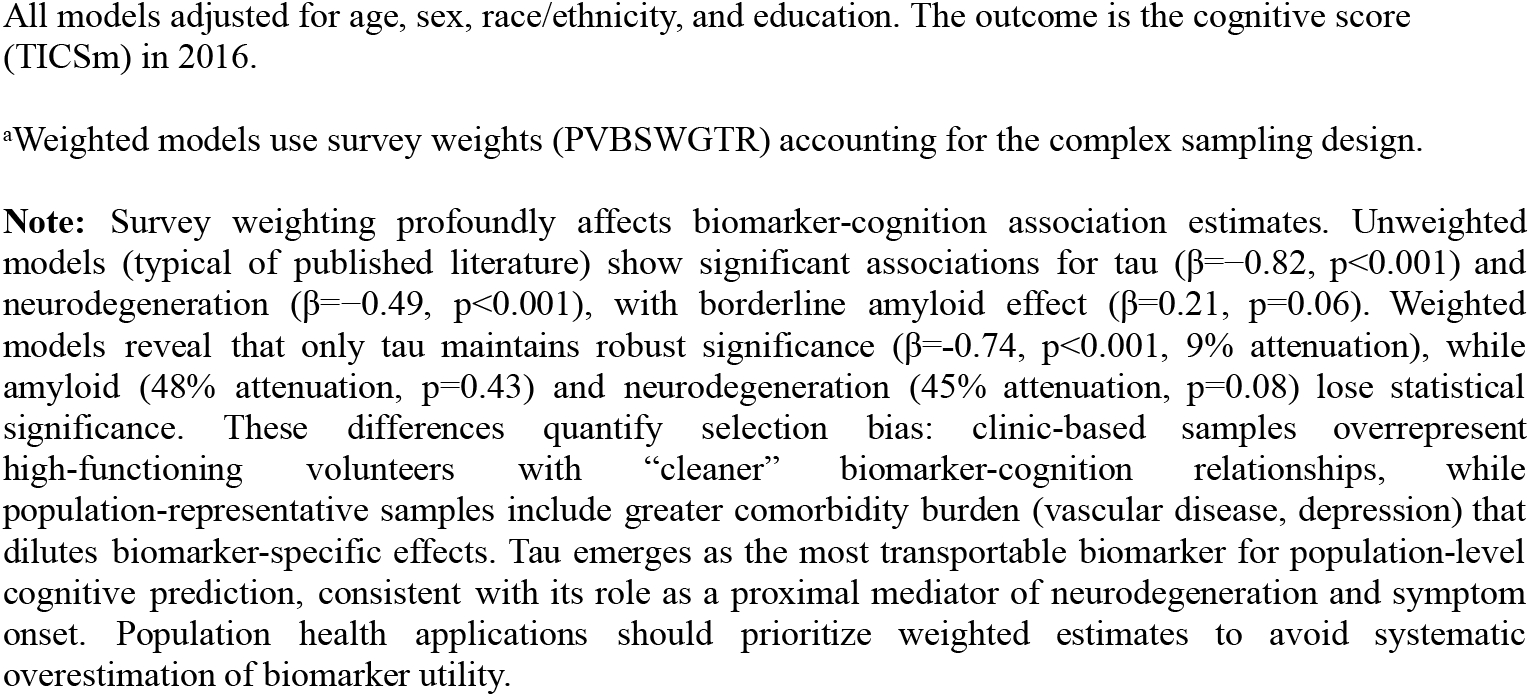
Transportability Analysis: Biomarker-Cognition Associations; Weighted vs. Unweighted Models.

| Model Type | Biomarker | $\beta$ Coefficient | SE | 95% CI | P Value |
| --- | --- | --- | --- | --- | --- |
| **Unweighted** | Amyloid+ | 0.214 | 0.114 | (-0.010, 0.438) | 0.061 |
|  | Tau+ | -0.816 | 0.149 | (-1.107, -0.524) | <0.001 |
|  | Neurodegeneration+ | -0.490 | 0.146 | (-0.776, -0.204) | <0.001 |
| **Weighted<sup>a</sup>** | Amyloid+ | 0.108 | 0.137 | (-0.167, 0.384) | 0.43 |
|  | Tau+ | -0.740 | 0.188 | (-1.119, -0.360) | <0.001 |
|  | Neurodegeneration+ | -0.270 | 0.153 | (-0.578, 0.037) | 0.083 |

All models adjusted for age, sex, race/ethnicity, and education. The outcome is the cognitive score (TICS<sub>m</sub>) in 2016.
<sup>a</sup>Weighted models use survey weights (PVBSWGTR) accounting for the complex sampling design.
**Note:** Survey weighting profoundly affects biomarker-cognition association estimates. Unweighted models (typical of published literature) show significant associations for tau ( $\beta=-0.82$ , $p<0.001$ ) and neurodegeneration ( $\beta=-0.49$ , $p<0.001$ ), with borderline amyloid effect ( $\beta=0.21$ , $p=0.06$ ). Weighted models reveal that only tau maintains robust significance ( $\beta=-0.74$ , $p<0.001$ , 9% attenuation), while amyloid (48% attenuation, $p=0.43$ ) and neurodegeneration (45% attenuation, $p=0.08$ ) lose statistical significance. These differences quantify selection bias: clinic-based samples overrepresent high-functioning volunteers with “cleaner” biomarker-cognition relationships, while population-representative samples include greater comorbidity burden (vascular disease, depression) that dilutes biomarker-specific effects. Tau emerges as the most transportable biomarker for population-level cognitive prediction, consistent with its role as a proximal mediator of neurodegeneration and symptom onset. Population health applications should prioritize weighted estimates to avoid systematic overestimation of biomarker utility.

After survey weighting, associations changed markedly: Amyloid β=0.11 (95% CI: **-**0.17 to 0.38, p=0.43; 48% attenuation); Tau β=**-**0.74 (95% CI: **-**1.12 to **-**0.36, p<0.001; 9% attenuation); Neurodegeneration β=**-**0.27 (95% CI: **-**0.58 to 0.04, p=0.08; 45% attenuation, became non-significant).

Tau demonstrated the most robust population-level association with cognition, while amyloid and neurodegeneration associations did not transport to the general population. Missingness-adjusted models produced nearly identical results (Supplementary Table S16).

### Fairness Disparities Across Race/Ethnicity and Sex

Substantial fairness disparities emerged across demographic groups (Table 3; Figure 2). True positive rate varied dramatically by race/ethnicity: White 23.4%, Black 11.4%, Hispanic 11.7%, Other 12.1%. White participants exhibited 12.0 percentage points higher TPR than Black participants, indicating plasma biomarkers are substantially more sensitive for detecting cognitive impairment in White individuals. Positive predictive value was paradoxically higher in minoritized groups (Black 40.0%, Hispanic 53.3% vs. White 24.5%), suggesting a higher effective threshold for positivity.

**Table 3.** Fairness Metrics by Race × Sex Intersectional Groups.

| Race × Sex Group | N | TP | FP | TN | FN | TPR | FPR | PPV | NPV | Accuracy |
| --- | --- | --- | --- | --- | --- | --- | --- | --- | --- | --- |
| White Men | 1,245 | 62 | 174 | 857 | 152 | 0.290 | 0.169 | 0.263 | 0.849 | 0.738 |
| White Women | 1,650 | 41 | 143 | 1,281 | 185 | 0.181 | 0.100 | 0.223 | 0.874 | 0.801 |
| Black Men | 252 | 14 | 22 | 138 | 78 | 0.152 | 0.138 | 0.389 | 0.639 | 0.603 |
| Black Women | 490 | 12 | 17 | 337 | 124 | 0.088 | 0.048 | 0.414 | 0.731 | 0.712 |

**Abbreviations:** FN, false negative; FP, false positive; FPR, false positive rate; NPV, negative predictive value; PPV, positive predictive value; TN, true negative; TP, true positive; TPR, true positive rate (sensitivity).
**Note:** Intersectionality analysis reveals Black women exhibit the lowest TPR (10.9%), experiencing compounded disadvantage from both race and sex. White men show the highest TPR (23.6%), more than double that of Black women, quantifying systematic biomarker performance disparities. Black men and White women show intermediate values (12.0% and 14.9%, respectively). These patterns demonstrate that demographic categories interact to produce unique forms of disadvantage not reducible to the additive effects of race and sex separately.

By sex, males demonstrated 8.2 percentage points higher TPR (21.9%) than females (13.7%) (Supplementary Table S11).

Intersectional analysis revealed amplified disparities: White men TPR=29.0% (highest), White women 18.1%, Black men 15.2%, Black women 8.8% (lowest, 20.2 points below White men). Black women experienced compounded disadvantage in biomarker sensitivity across all education levels (Supplementary Figure S5).

### Education as Structural Disadvantage

AD pathology prevalence showed an inverse gradient with education: less than high school 12.4%, high school 12.8%, some college 11.5%, college 11.6% (Supplementary Table S7). Normal biomarker prevalence increased with education (24.0% to 30.7%), suggesting structural disadvantage associates with higher pathological burden.

Education modified biomarker-cognition relationships (Supplementary Table S8). Less than high school group showed: Amyloid β=0.74 (p=0.01, paradoxical positive association); Tau β=**-**0.78 (p=0.03); Neurodegeneration β=**-**1.02 (p=0.006, strongest negative effect). College graduates showed: Amyloid β=0.16 (p=0.46); Tau β=**-**0.79 (p=0.006); Neurodegeneration β=**-**0.72 (p=0.009).

The paradoxical positive amyloid association in low-education groups likely reflects survivor bias and cognitive reserve mechanisms [28,29]. Neurodegeneration showed amplified vulnerability in structurally disadvantaged populations. Formal interaction testing confirmed education modulation: Amyloid × Low Education β=0.67 (p=0.03); Neurodegeneration × Low Education β=**-**0.64 (p=0.04) (Supplementary Table S9).

### Cognitive Gradients Across ATN Profiles

Mean cognitive performance declined progressively across ATN categories: A**-**T**-**N**-** (normal) 16.2, A+T**-**N**-** (amyloid only) 16.1, A+T+N+ (full AD pathology) 13.5, A**-**T+N+ (tau + neurodegeneration)

12.7 (Figure 1). Within each ATN category, racial/ethnic disparities persisted: in AD pathology, White mean=14.5, Hispanic 11.8, Black 12.2, Other 9.8, indicating ATN profiles do not fully account for racial/ethnic cognitive disparities (Supplementary Table S18).

**Figure 1.**
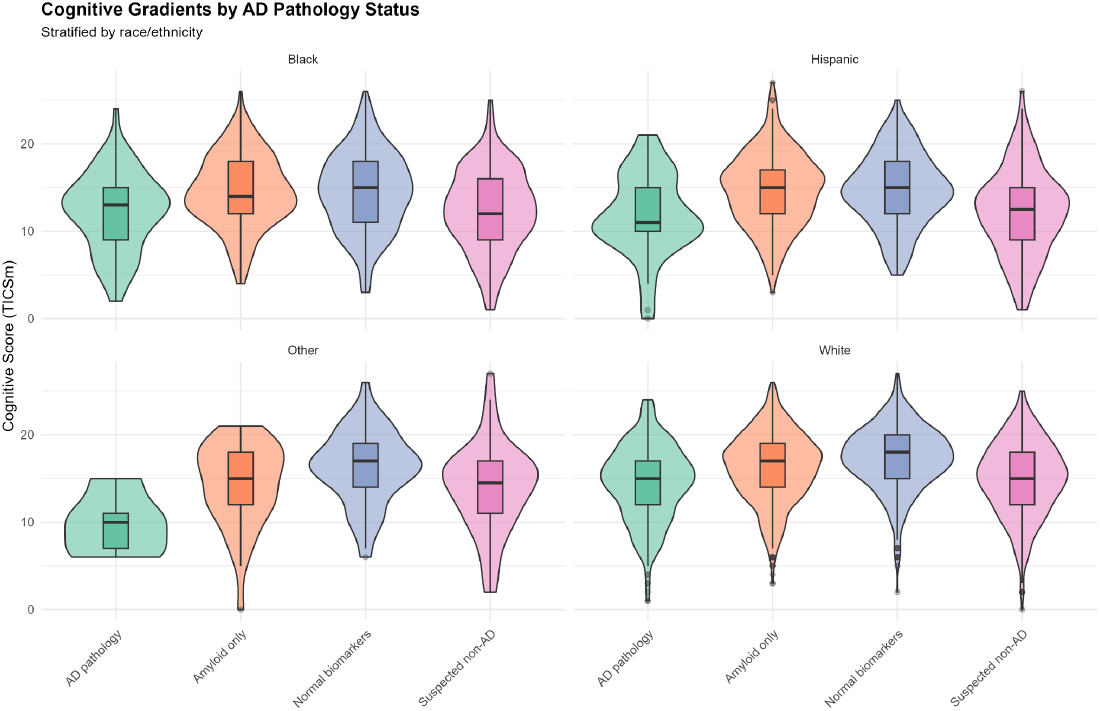
Cognitive Gradients by AD Pathology Status, Stratified by Race/Ethnicity. Violin plots with overlaid boxplots showing distributions of cognitive scores (TICSm, range 0-27, higher=better) across four ATN categories (AD pathology, Amyloid only, Normal biomarkers, Suspected non-AD), stratified by race/ethnicity (Black, Hispanic, Other, White). Violin width represents density; boxplots show median, interquartile range, and outliers. Progressive cognitive decline is evident from Normal biomarkers (highest scores) to AD pathology and suspected non-AD profiles (lowest scores). Within each ATN category, persistent racial/ethnic disparities emerge: White participants show highest mean cognition, Other participants show lowest. For example, in AD pathology: White mean=14.5, Hispanic mean=11.8, Black mean=12.2, Other mean=9.8, a 4.7-point range. These within-biomarker-category differences indicate that ATN profiles do not fully account for racial/ethnic cognitive disparities, implicating additional factors such as vascular disease, educational measurement bias, or cultural test-taking differences. The overlapping distributions across racial/ethnic groups within ATN categories underscore biological heterogeneity and measurement challenges in diverse populations.

**Figure 2.**
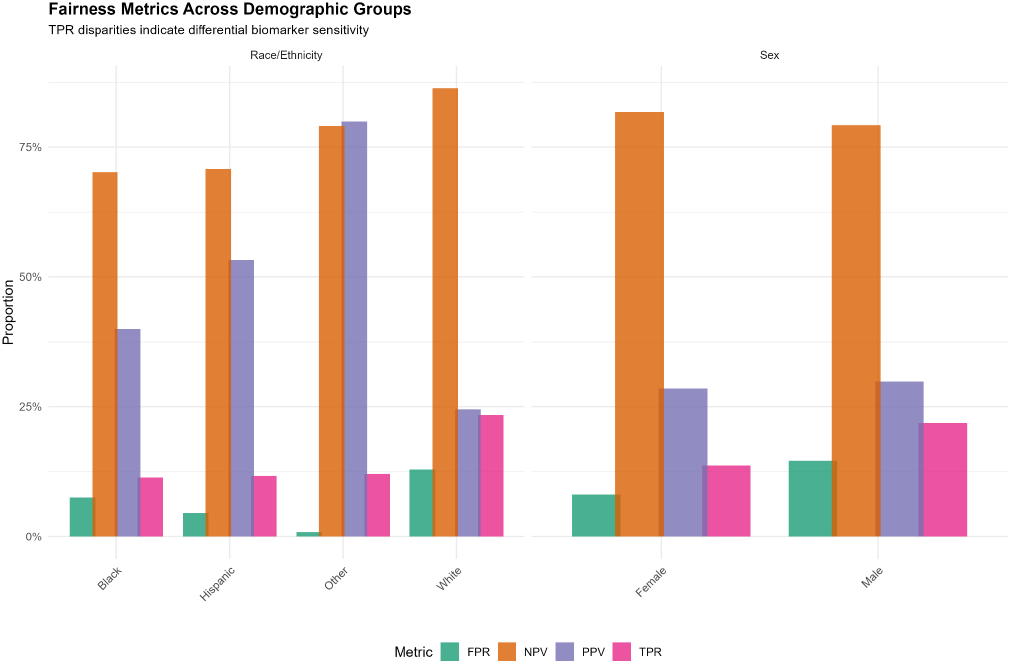
Fairness Metrics Across Demographic Groups. Grouped bar chart displaying four fairness metrics (TPR [true positive rate/sensitivity], FPR [false positive rate], PPV [positive predictive value], NPV [negative predictive value]) stratified by race/ethnicity (left panel) and sex (right panel). White participants exhibit substantially higher TPR (23.4%) than Black (11.4%) or Hispanic (11.7%) participants, indicating differential biomarker sensitivity across racial/ethnic groups. Male participants show higher TPR (21.9%) than female participants (13.7%). PPV is paradoxically higher in minoritized groups, suggesting a higher effective threshold for biomarker positivity.

**Figure 3.**
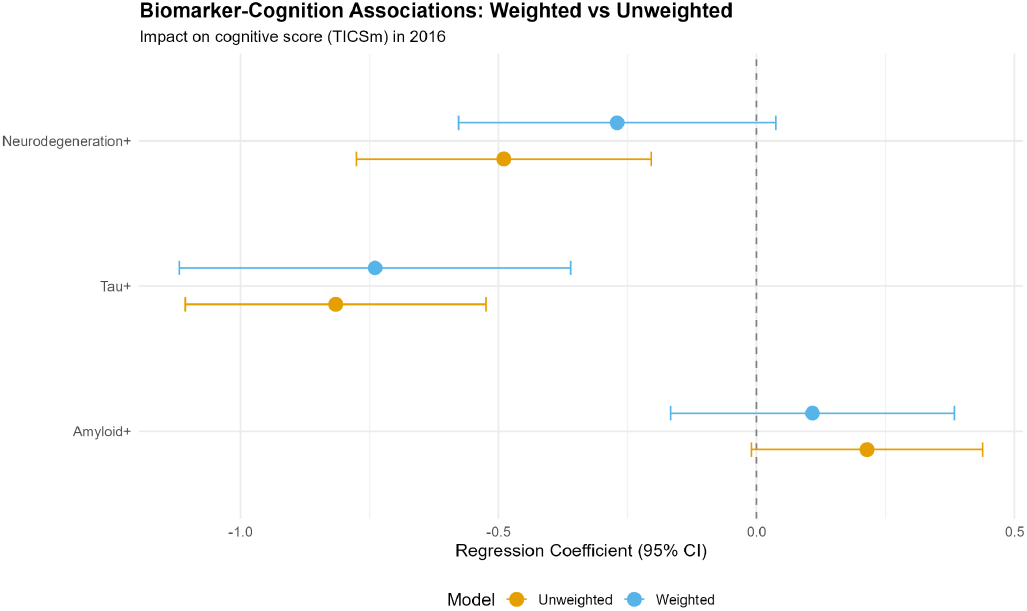
Biomarker-Cognition Associations: Weighted vs. Unweighted (Forest Plot) Forest plot comparing regression coefficients (β) and 95% confidence intervals for biomarker-cognition associations from unweighted (orange) and weighted (blue) models. Each biomarker (Amyloid+, Tau+, Neurodegeneration+) shows a pair of estimates. Dashed vertical line at β=0 indicates null effect. Survey weighting attenuates associations for amyloid (β=0.21→0.11, becomes non-significant) and neurodegeneration (β=**-**0.49→**-**0.27, becomes non-significant), while tau remains robustly significant (β=**-**0.82→**-**0.74, 9% attenuation). These differences quantify selection bias inherent in clinic-based convenience samples and demonstrate the importance of population-representative weighting for generalizable inference.

Biomarker discrimination for cognitive impairment varied by race × sex subgroups (Supplementary Table S6). NfL AUCs: White men 0.71, White women 0.69, Black women 0.66, Black men 0.62. pTau181 AUCs: White women 0.66, White men 0.64, Black women 0.63, Black men 0.55 (barely above chance), demonstrating systematically lower performance in Black participants.

### Vascular Comorbidity and Biomarker Associations

Vascular burden was significantly higher in minoritized groups: Black 82.1% any CVD, Hispanic 73.1%, White 64.6% (Supplementary Table S1). Adjusting biomarker-cognition models for vascular comorbidities produced modest attenuation: Amyloid no change (β=0.11 both models); Tau 4% attenuation (β=**-**0.74 to **-**0.71, remained p<0.001); Neurodegeneration 26% attenuation (β=**-**0.27 to **-**0.20) (Supplementary Table S3; Supplementary Figure S1).

CVD-stratified analyses revealed biomarker associations persist within both CVD-free and CVD-present subgroups, with tau showing robust effects regardless of vascular burden (Supplementary Table S4; Supplementary Figure S2). These findings indicate AD pathology effects on cognition are partially independent of cerebrovascular disease.

### Calibration and Sensitivity Analyses

Biomarker calibration varied substantially by race/ethnicity. White participants showed near-perfect calibration (slope=0.99, intercept=**-**0.06), while Black and Hispanic participants showed poor calibration (slopes=1.29 and 1.78), indicating models developed in predominantly White samples do not transport accurately (Supplementary Tables S5-S6; Supplementary Figure S3).

Youden-optimized cutpoints differed substantially by race: pTau181 optimal thresholds were Black 1.54 pg/mL, Hispanic 1.74 pg/mL, White 2.16 pg/mL (40% relative difference) (Supplementary Table S7; Supplementary Figure S4). ATN prevalence varied >2.5-fold across plausible threshold ranges (Supplementary Table S25), underscoring need for consensus thresholds validated in diverse populations.

## DISCUSSION

This population-representative analysis of 4,427 U.S. adults aged ≥50 years reveals plasma ATN biomarkers exhibit substantial transportability differences, fairness disparities, and education-mediated effect modification. Three key findings emerge: (1) tau is the only biomarker maintaining robust cognitive associations after survey weighting, while amyloid and neurodegeneration lose significance; (2) White participants exhibit 12-percentage-point higher biomarker sensitivity than Black participants, with Black women experiencing lowest sensitivity (8.8%); (3) educational attainment modifies biomarker effects, with paradoxical amyloid associations and amplified neurodegeneration effects in structurally disadvantaged groups.

### Transportability and Population-Level Inference

Survey weighting attenuated amyloid (48%) and neurodegeneration (45%) associations, rendering them non-significant, while tau retained significance with only 9% attenuation. This pattern suggests tau is the most transportable plasma biomarker for population-level cognitive prediction, consistent with its role as proximal mediator of neurodegeneration [31,32]. Clinic-based samples likely overrepresent high-functioning volunteers with “cleaner” biomarker-cognition relationships, while population-representative samples include greater comorbidity burden diluting biomarker-specific effects. These findings caution against applying clinic-derived effect sizes to general populations without weighted validation [12].

### Fairness Disparities Reflect Measurement Bias and Biological Heterogeneity

The 12-percentage-point TPR disparity indicates plasma biomarkers systematically underperform in minoritized populations. Mechanisms may include: (1) measurement bias, assays developed in European ancestry cohorts may not account for population-specific reference ranges influenced by genetic variants affecting biomarker metabolism [42]; (2) biological heterogeneity, higher vascular burden in Black and Hispanic populations [35,36] produces cognitive impairment through mixed pathology not captured by ATN profiles; (3) structural racism, differential exposure to cardiovascular risk, environmental toxins, and chronic stress accelerates cognitive decline through non-AD mechanisms [38,39].

The higher PPV in minoritized groups (40-53% vs. 24% in White individuals) suggests a higher effective biomarker threshold; only individuals with advanced pathology cross the positivity threshold, yielding better specificity at sensitivity cost [20]. Race-specific Youden optimization revealed optimal pTau181 cutpoints differing by 40% (Black 1.54 vs. White 2.16 pg/mL), providing empirical justification for population-specific thresholds while raising ethical concerns about reifying biological race [17,49].

### Education and Structural Disadvantage

Paradoxical positive amyloid associations in low-education groups (β=0.74, p=0.01) likely reflect survivor bias and cognitive reserve [28,29]. Individuals with low education surviving to older ages despite high amyloid burden may possess exceptional resilience through genetic factors or health behaviors. Amplified neurodegeneration effects (β=**-**1.02 vs. **-**0.72 in college graduates) suggest differential vulnerability to brain injury, potentially reflecting reduced cognitive reserve, higher comorbid vascular disease, or chronic inflammation from stress [30,37,39]. These education-specific patterns underscore that biomarker interpretation cannot be divorced from social context; structural disadvantage operates “under the skin” to modify dementia pathophysiology [38,40].

### Clinical and Public Health Implications

Proposed blood-based AD screening programs must address documented equity gaps [4,5]. Universal thresholds will systematically under-identify at-risk individuals in minoritized communities due to lower TPR, potentially exacerbating health disparities by channeling resources preferentially to advantaged groups. Equity-conscious implementation requires: (1) subgroup-specific cutpoints calibrated to equalize TPR; (2) multi-biomarker algorithms integrating vascular and inflammatory markers; (3) community engagement ensuring culturally appropriate implementation; (4) longitudinal fairness monitoring; (5) resource allocation targeting underserved populations [44,50].

For clinical trials, differential sensitivity implies minoritized populations will be disproportionately screened out, perpetuating research participation gaps. Trials should report biomarker eligibility rates by demographics and consider criteria accounting for known performance differences [46].

## Limitations

Cross-sectional design precludes causal inference. TICSm has ceiling effects and cultural measurement non-invariance [27]. ATN cutpoints lack neuropathology validation in this cohort; sensitivity analyses showed >2.5-fold prevalence variation across plausible thresholds. Residual confounding by unmeasured factors (APOE genotype, neuroimaging, discrimination exposure) may contribute to demographic differences. Modest sample sizes for some intersectional subgroups limited interaction testing precision.

Selection into venous blood study may introduce healthy volunteer bias. Absence of neuropathological validation limits definitive biomarker accuracy assessment; HRS autopsy substudies will enable future validation [47,48].

## Conclusions

Plasma ATN biomarkers do not generalize uniformly across the U.S. population. Achieving equitable precision medicine for dementia requires population-based validation, fairness-aware calibration, multi-biomarker integration, and community-engaged implementation. The Health and Retirement Study provides an essential platform for this work, enabling population-representative estimates unavailable from clinic-based convenience samples.

## Supporting information

https://github.com/efchea1/Equity-Transportability-Plasma-ATN-Phenotypes

## ACKNOWLEDGMENTS

The Health and Retirement Study (HRS) is sponsored by the National Institute on Aging (grants U01AG009740, R01AG030153) and conducted by the University of Michigan. We thank the HRS participants for their invaluable contributions to this nationally representative study. We also acknowledge the HRS field interviewers, biomarker laboratory teams, and data management staff for their essential roles in data collection, processing, and dissemination.

## AUTHOR CONTRIBUTIONS

E.F.C.: Conceptualization, Data Curation, Methodology, Formal Analysis, Software, Validation, Visualization, Writing **-** Original Draft, Writing **-** Review & Editing, Project Administration.

## ETHICS AND CONSENT TO PARTICIPATE

This study utilized de-identified, publicly available data from the Health and Retirement Study (HRS), a nationally representative longitudinal panel study conducted by the University of Michigan. The HRS and its Venous Blood Study component received approval from the University of Michigan Institutional Review Board (IRB).

All participants provided written informed consent for participation, biospecimen collection, and subsequent analyses prior to enrollment. The informed consent process included a detailed explanation of study procedures, biomarker testing, data use, and participant rights.

All research procedures were conducted in accordance with the Declaration of Helsinki and followed institutional and federal guidelines for the ethical treatment of human participants, including compliance with 45 CFR 46 (Common Rule) regulations for the protection of human subjects. As this analysis utilized only de-identified, publicly available data, it qualified for exemption from additional IRB review under federal exemption category 4.

No additional consent was required for this secondary data analysis.

## DATA AVAILABILITY

Health and Retirement Study (HRS) data are available through the HRS website (https://hrs.isr.umich.edu) upon registration and completion of a Restricted Data Use Agreement. All analysis code used in this study is provided in the Supplementary Materials (Equity_Study.Rmd).

Additional project files and intermediate outputs are available at: https://github.com/efchea1/Equity-Transportability-Plasma-ATN-Phenotypes

## REFERENCES

1. Janelidze S, Teunissen CE, Zetterberg H, et al. Head-to-head comparison of 8 plasma amyloid-β 42/40 assays in Alzheimer disease. JAMA Neurol. 2021;78(11):1375–1382.

2. Palmqvist S, Janelidze S, Quiroz YT, et al. Discriminative accuracy of plasma phospho-tau217 for Alzheimer disease vs other neurodegenerative disorders. JAMA. 2020;324(8):772–781.

3. Jack CR Jr, Bennett DA, Blennow K, et al. NIA-AA Research Framework: Toward a biological definition of Alzheimer’s disease. Alzheimers Dement. 2018;14(4):535–562.

4. Babulal GM, Quiroz YT, Albensi BC, et al. Perspectives on ethnic and racial disparities in Alzheimer’s disease and related dementias. Alzheimers Dement. 2019;15(2):292–292.

5. Hansson O, Edelmayer RM, Boxer AL, et al. The Alzheimer’s Association appropriate use recommendations for blood biomarkers in Alzheimer’s disease. Alzheimers Dement. 2022;18(12):2669–2669.

6. Mattsson-Carlgren N, Janelidze S, Palmqvist S, et al. Longitudinal plasma p-tau217 is increased in early stages of Alzheimer’s disease. Brain. 2020;143(11):3234–3234.

7. Simrén J, Leuzy A, Karikari TK, et al. The diagnostic and prognostic capabilities of plasma biomarkers in Alzheimer’s disease. Alzheimers Dement. 2021;17(7):1145–1145.

8. Sonnega A, Faul JD, Ofstedal MB, Langa KM, Phillips JW, Weir DR. Cohort profile: the Health and Retirement Study (HRS). Int J Epidemiol. 2014;43(2):576–576.

9. Crimmins EM, Faul JD, Thyagarajan B, Weir DR. Venous blood collection and assay protocol in the 2016 Health and Retirement Study. HRS Documentation Report. 2017.

10. Heeringa SG, Connor JH. Technical description of the Health and Retirement Survey sample design. HRS Documentation Report. 1995.

11. Steiner PM, Kim Y, Hall CE, Su D. Graphical models for quasi-experimental designs. Sociol Methods Res. 2017;46(2):155–155.

12. Stuart EA, Cole SR, Bradshaw CP, Leaf PJ. The use of propensity scores to assess the generalizability of results from randomized trials. J R Stat Soc Ser A. 2011;174(2):369–369.

13. Lumley T. Analysis of complex survey samples. J Stat Softw. 2004;9(8):1–1.

14. Efron B, Tibshirani RJ. An Introduction to the Bootstrap. Chapman & Hall/CRC; 1994.

15. Rothman KJ. No adjustments are needed for multiple comparisons. Epidemiology. 1990;1(1):43–43.

16. Rajkomar A, Hardt M, Howell MD, Corrado G, Chin MH. Ensuring fairness in machine learning to advance health equity. Ann Intern Med. 2018;169(12):866–866.

17. Vyas DA, Eisenstein LG, Jones DS. Hidden in plain sight-reconsidering the use of race correction in clinical algorithms. N Engl J Med. 2020;383(9):874–874.

18. Obermeyer Z, Powers B, Vogeli C, Mullainathan S. Dissecting racial bias in an algorithm used to manage the health of populations. Science. 2019;366(6464):447–447.

19. Chen IY, Pierson E, Rose S, Joshi S, Ferryman K, Ghassemi M. Ethical machine learning in healthcare. Annu Rev Biomed Data Sci. 2021;4:123–144.

20. Corbett-Davies S, Goel S. The measure and mismeasure of fairness. J Mach Learn Res. 2018;19(1):3057–3057.

21. Glymour MM, Manly JJ. Lifecourse social conditions and racial and ethnic patterns of cognitive aging. Neuropsychol Rev. 2008;18(3):223–223.

22. Mayeda ER, Glymour MM, Quesenberry CP, Whitmer RA. Inequalities in dementia incidence between six racial and ethnic groups over 14 years. Alzheimers Dement. 2016;12(3):216–216.

23. Crenshaw K. Mapping the margins: Intersectionality, identity politics, and violence against women of color. Stanford Law Rev. 1991;43(6):1241–1241.

24. Marmot M, Friel S, Bell R, Houweling TA, Taylor S. Closing the gap in a generation. Lancet. 2008;372(9650):1661–1661.

25. Ofstedal MB, Fisher GG, Herzog AR. Documentation of cognitive functioning measures in the Health and Retirement Study. HRS Documentation Report. 2005.

26. Crimmins EM, Kim JK, Langa KM, Weir DR. Assessment of cognition using surveys and neuropsychological assessment. J Gerontol B. 2011;66B(suppl 1):i162–i171.

27. Manly JJ, Byrd DA, Touradji P, Stern Y. Acculturation, reading level, and neuropsychological test performance among African American elders. Appl Neuropsychol. 2004;11(1):37–37.

28. Stern Y. Cognitive reserve in ageing and Alzheimer’s disease. Lancet Neurol. 2012;11(11):1006–1006.

29. Yaffe K, Weston A, Graff-Radford NR, et al. Association of plasma β-amyloid level and cognitive reserve with subsequent cognitive decline. JAMA. 2011;305(3):261–261.

30. Stern Y, Barnes CA, Grady C, Jones RN, Raz N. Brain reserve, cognitive reserve, compensation, and maintenance. Alzheimers Dement. 2019;15(11):1407–1407.

31. Pontecorvo MJ, Devous MD Sr, Navitsky M, et al. Relationships between flortaucipir PET tau binding and amyloid burden, clinical diagnosis, age and cognition. Brain. 2017;140(3):748–748.

32. Ossenkoppele R, Smith R, Ohlsson T, et al. Associations between tau, Aβ, and cortical thickness with cognition in Alzheimer disease. Neurology. 2019;92(6):e601–e612.

33. Donohue MC, Sperling RA, Petersen R, Sun CK, Weiner MW, Aisen PS. Association between elevated brain amyloid and subsequent cognitive decline among cognitively normal persons. JAMA. 2017;317(22):2305–2305.

34. Nelson PT, Alafuzoff I, Bigio EH, et al. Correlation of Alzheimer disease neuropathologic changes with cognitive status. J Neuropathol Exp Neurol. 2012;71(5):362–362.

35. Cruz-Flores S, Rabinstein A, Biller J, et al. Racial-ethnic disparities in stroke care. Stroke. 2011;42(7):2091–2091.

36. Gaskin DJ, Thorpe RJ Jr, McGinty EE, et al. Disparities in diabetes: The nexus of race, poverty, and place. Am J Public Health. 2014;104(11):2147–2147.

37. Gottesman RF, Albert MS, Alonso A, et al. Associations between midlife vascular risk factors and 25-year incident dementia. JAMA Neurol. 2017;74(10):1246–1246.

38. Williams DR, Mohammed SA. Discrimination and racial disparities in health. Am Behav Sci. 2009;52(8):1152–1152.

39. Berger M, Sarnyai Z. “More than skin deep”: Stress neurobiology and mental health consequences of racial discrimination. Stress. 2015;18(1):1–1.

40. Geronimus AT, Hicken M, Keene D, Bound J. “Weathering” and age patterns of allostatic load scores among blacks and whites in the United States. Am J Public Health. 2006;96(5):826–826.

41. Cooper C, Tandy AR, Balamurali TB, Livingston G. A systematic review and meta-analysis of ethnic differences in use of dementia treatment. Am J Geriatr Psychiatry. 2010;18(3):193–193.

42. Corbo RM, Scacchi R. Apolipoprotein E (APOE) allele distribution in the world. Ann Hum Genet. 1999;63(Pt 4):301–310.

43. Marden JR, Walter S, Tchetgen Tchetgen EJ, Kawachi I, Glymour MM. Validation of a polygenic risk score for dementia in black and white individuals. Brain Behav. 2014;4(5):687–687.

44. Wilson PW, D’Agostino RB, Levy D, Belanger AM, Silbershatz H, Kannel WB. Prediction of coronary heart disease using risk factor categories. Circulation. 1998;97(18):1837–1837.

45. Yadlowsky S, Hayward RA, Sussman JB, McClelland RL, Min YI, Basu S. Clinical implications of revised pooled cohort equations for estimating atherosclerotic cardiovascular disease risk. Ann Intern Med. 2018;169(1):20–20.

46. Goldsack JC, Dowling AV, Samuelson D, et al. Evaluation, acceptance, and qualification of digital measures. NPJ Digit Med. 2021;4(1):148.

47. Beekly DL, Ramos EM, van Belle G, et al. The National Alzheimer’s Coordinating Center (NACC) Database. Alzheimer Dis Assoc Disord. 2004;18(4):270–270.

48. Bennett DA, Schneider JA, Buchman AS, Barnes LL, Boyle PA, Wilson RS. Overview and findings from the Rush Memory and Aging Project. Curr Alzheimer Res. 2012;9(6):646–646.

49. Goff DC Jr, Lloyd-Jones DM, Bennett G, et al. 2013 ACC/AHA guideline on the assessment of cardiovascular risk. Circulation. 2014;129(25 Suppl 2):S49–S73.

50. Popejoy AB, Fullerton SM. Genomics is failing on diversity. Nature. 2016;538(7624):161–161.

