## Supplementary material for "Equity and Transportability of Plasma ATN Phenotypes in a Population-Representative U.S. Aging Cohort": https://github.com/efchea1/Equity-Transportability-Plasma-ATN-Phenotypes

### SUPPLEMENTARY MATERIALS

### SUPPLEMENTARY METHODS

**Survey Weight Construction and Application**

HRS employs a complex multistage probability sampling design with stratification by geography and race/ethnicity, clustering within primary sampling units, and oversampling of Black and Hispanic individuals. Person-level survey weights (PVBSWGTR) adjust for: (1) base sampling probability; (2) non-response at household and individual levels; (3) post-stratification to Current Population Survey marginal totals for age, sex, and race/ethnicity.

We applied survey weights using the R survey package [20], specifying:

- **Weights:** PVBSWGTR
- **Strata:** STRATUM
- **Primary sampling units:** SECU
- **Nesting:** TRUE (to accommodate certainty PSUs)

All prevalence estimates, means, regression coefficients, and confidence intervals presented in the main manuscript reflect survey-weighted analyses unless explicitly noted as "unweighted."

#### Inverse Probability Weighting for Missingness

To assess whether differential missingness biased estimates, we constructed inverse probability weights (IPW) as follows:

1. Fitted logistic regression predicting complete-case status (all biomarkers and cognition observed) from age, sex, race/ethnicity, and education.
2. Predicted probability of being a complete case for all individuals: $\widehat{p}_i = P(Complete \mid X_i$
3. Calculated IPW: $w_i^{IPW} = \frac{1}{\widehat{p}_i}$ for complete cases, 0 otherwise.
4. Combined with survey weights: $w_i^{combined} = w_i^{survey} \times w_i^{IPW}$
5. Truncated at the 99th percentile to reduce extreme weight influence.

Models using combined weights produced nearly identical results to standard survey-weighted models (Table 16), indicating that missingness was missing at random conditional on observed covariates.

#### Bootstrap Internal Validation

To assess internal validity and quantify sampling variability, we conducted 1,000 bootstrap resamples with replacement from the complete-case analytic sample. For each resample, we calculated:

- ATN prevalence for all eight profiles
- Regression coefficients for biomarker-cognition associations

Bootstrap percentile confidence intervals (2.5th and 97.5th percentiles) are reported in Tables 20-21. Narrow intervals confirm adequate statistical power and stable estimates.

#### Youden Index Optimization

For each racial/ethnic group separately, we identified optimal biomarker cutpoints maximizing the Youden index:

$$J = Sensitivity + Specificity - 1$$

using the cutpointr R package. Outcome was cognitive impairment (CIND or dementia). Results are presented in Table S7 and Figure S4.

These race-specific optima provide empirical justification for considering population-specific thresholds, though ethical and practical considerations around race-specific algorithms must be weighed carefully.

### SUPPLEMENTAL STRENGTHS AND LIMITATIONS

#### Strengths

This study's primary strength is its use of the Health and Retirement Study, the only nationally representative U.S. aging cohort with plasma AD biomarker data. The probability sampling design, oversampling of Black and Hispanic individuals, and application of survey weights enable population-level inference unavailable from clinic-based cohorts. The large sample size (n=4,427) provides adequate power for subgroup analyses while maintaining meaningful representation of structurally disadvantaged groups. Methodological rigor is enhanced through complex survey weighting, inverse probability weighting for missingness, bootstrap internal validation, calibration analyses, and extensive sensitivity analyses across biomarker thresholds. The integration of transportability, fairness, and structural disadvantage frameworks represents a novel contribution to the AD biomarker literature.

#### Limitations

Several methodological limitations warrant consideration. First, the cross-sectional design precludes causal inference regarding the temporal ordering of biomarker changes and cognitive decline. While biomarker-cognition associations are interpreted as reflecting underlying pathology, reverse causation or bidirectional processes cannot be excluded. Future waves of HRS biomarker data will enable longitudinal modeling of biomarker trajectories.

Second, cognitive outcomes were reconstructed from TICSm items across 2016-2020. Although validated for population studies, TICSm is a brief screening tool with ceiling effects, limited domain coverage, and cultural measurement non-invariance. Proxy and self-reported cognition were collapsed following HRS guidelines, but proxy assessments may introduce systematic downward bias. Wave-specific differences in interview mode, particularly during the COVID-19 pandemic, may also affect measurement consistency.

Third, ATN cutpoints (Aβ42/40 $<0.063$, pTau181 $>2.5$ pg/mL, NfL $>20$ pg/mL) were derived from predominantly European-ancestry clinical cohorts and may not be optimal for diverse populations. Sensitivity analyses revealed $>2.5$-fold variation in AD pathology prevalence across plausible thresholds. Race-specific Youden-optimized cutpoints differed by up to 40%, highlighting the need for neuropathology-validated thresholds in heterogeneous populations.

Fourth, residual confounding remains possible. APOE ε4 genotype, neuroimaging markers (e.g., white matter hyperintensities), chronic stress biomarkers, discrimination exposure, and neighborhood disadvantage were unavailable but likely contribute to demographic differences in biomarker performance. Incorporating these factors in future analyses will strengthen causal inference.

Fifth, some intersectional subgroups, particularly Black men and individuals classified as “Other” race, had modest sample sizes, limiting precision for interaction models and ROC analyses. Confidence intervals for subgroup AUCs were wide, especially for pTau181 in Black men. Pooling across cohorts or leveraging future HRS biomarker waves will improve precision.

Sixth, selection into the Venous Blood Study may introduce healthy volunteer bias. Participation required in-person interviews and venipuncture, potentially excluding individuals with severe impairment, mobility limitations, or medical contraindications. Although survey weighting mitigates this bias, underrepresentation of the most impaired individuals may attenuate biomarker–cognition associations and influence fairness metrics.

Seventh, biomarker analytical variability across assay batches, plates, or operators was not fully characterized in the available data. Pre-analytical factors such as hemolysis, lipemia, and freeze-thaw cycles may vary across demographic groups and introduce differential measurement error. Rigorous quality control procedures, including blinded duplicates and calibration standards, will be essential for multi-site implementation.

Finally, the absence of neuropathological validation in this cohort limits definitive assessment of biomarker accuracy. While cognitive impairment serves as a clinically meaningful outcome, plasma biomarkers should ultimately be validated against autopsy diagnoses. Ongoing HRS autopsy substudies will enable pathology-anchored validation in future work.

### SUPPLEMENTARY RESULTS

#### Vascular Comorbidity Prevalence by Race/Ethnicity and Sex

Vascular comorbidity burden was substantially higher in minoritized groups (Table S1). Black participants exhibited the highest prevalence of hypertension (78.2%), diabetes (35.7%), and any CVD (82.1%).

Hispanic participants showed intermediate prevalence (hypertension 64.7%, diabetes 38.1%, any CVD 73.1%). White participants had the lowest burden (hypertension 59.4%, diabetes 22.5%, any CVD 64.6%).

Within racial/ethnic groups, sex differences were modest, though Black men showed slightly higher CVD prevalence than Black women (83.7% vs. 81.3%; Table S2).

#### Impact of Vascular Adjustment on Biomarker-Cognition Associations

Adjusting for vascular comorbidities (hypertension, diabetes, stroke) modestly attenuated biomarker-cognition associations (Table S3; Figure S1):

- **Amyloid:** No change (β=0.11, both models)
- **Tau:** 4% attenuation (β=-0.74 → -0.71), remained highly significant (p<0.001)
- **Neurodegeneration:** 26% attenuation (β=−0.27 → -0.20), remained non-significant

These results indicate that observed biomarker-cognition relationships are not fully explained by vascular confounding, supporting partially independent AD pathology effects.

#### CVD-Stratified Biomarker Associations

Stratifying by CVD presence revealed that biomarker associations persist within both CVD-free and CVD-present subgroups (Table S4; Figure S2):

**No CVD (n=1,341):**

- Tau: β=-1.08, p<0.001
- Neurodegeneration: β=-0.29, p=0.28
- Amyloid: β=0.03, p=0.87

**Any CVD (n=2,955):**

- Tau: β=-0.72, p<0.001
- Neurodegeneration: β=-0.51, p=0.004
- Amyloid: β=0.27, p=0.05

Tau demonstrated robust associations regardless of vascular burden, while neurodegeneration showed stronger effects in the CVD-present group, potentially reflecting combined AD and vascular contributions to brain injury.

#### Calibration Analysis Details

Calibration was assessed by dividing participants into risk deciles based on predicted probability of cognitive impairment from logistic regression (A+T+N biomarkers as predictors). Within each decile and racial/ethnic group, we calculated the mean predicted risk and observed cognitive impairment rate (Table S5).

Perfect calibration corresponds to points on the 45-degree diagonal (predicted = observed). Deviations indicate systematic over- or under-prediction. Calibration statistics (Table S6) quantify model fit:

- **Calibration slope:** 1.0 indicates perfect calibration; >1 indicates overly narrow prediction intervals; <1 indicates overly wide.
- **Calibration intercept:** 0 indicates no systematic bias; <0 indicates overprediction; >0 indicates underprediction.

White participants showed near-perfect calibration (slope=0.99, intercept=-0.06), while Black and Hispanic participants showed poor calibration (slopes=1.29 and 1.78, intercepts=0.06 and −0.02), indicating that models developed in predominantly White samples do not transport accurately to minoritized populations (Figure S3).

#### Race-Specific Optimal Cutpoints

Youden-optimized cutpoints differed substantially by race (Table S7; Figure S4):

**pTau181:**

- Black: 1.54 pg/mL
- Hispanic: 1.74 pg/mL
- White: 2.16 pg/mL

**NfL:**

- Black: 17.6 pg/mL
- Hispanic: 21.6 pg/mL
- White: 24.0 pg/mL

**GFAP:**

- Black: 119.5 pg/mL
- Hispanic: 89.0 pg/mL
- White: 102.9 pg/mL

These differences (up to 40% relative) suggest that universal cutpoints may systematically misclassify minoritized individuals, contributing to observed fairness disparities. However, implementing race-specific thresholds raises ethical concerns about reifying biological race concepts and requires careful consideration of potential harms.

#### Fairness Landscape Across Education and Intersectional Groups

The fairness landscape heatmap (Figure S5) visualizes TPR across race × sex × education intersectional groups. Black women show persistently low TPR across all education levels, with particularly striking deficits in higher education strata:

- **Black women, less than high school:** TPR=7.9%
- **Black women, high school:** TPR=9.5%
- **Black women, some college:** TPR=5.3%
- **Black women, college+:** TPR=16.7%

In contrast, White men show higher TPR across education levels:

- **White men, less than high school:** TPR=24.1%
- **White men, high school:** TPR=35.3%
- **White men, some college:** TPR=15.4%
- **White men, college+:** TPR=36.1%

These patterns demonstrate that educational attainment does not fully mitigate race/sex-based biomarker performance gaps, consistent with intersectionality theory and research on diminishing returns to socioeconomic mobility for Black women [58].

### Supplementary Discussion

#### Comparison to ADNI and Other Clinical Cohorts

Our HRS sample differs markedly from ADNI participants along multiple dimensions (Table S9). ADNI is predominantly White (>90%), highly educated (>60% college+), and older (mean age 75 vs. 68.3 in HRS). These demographic differences have profound implications for biomarker performance estimates.

ADNI-derived cutpoints and effect sizes may not transport to general populations. For example, ADNI reports amyloid-cognition associations of $\beta \approx -1.0$ [32], substantially larger than our weighted estimate ($\beta = 0.11$, non-significant). This discrepancy likely reflects ADNI's enrichment for "pure" AD pathology, higher baseline cognition, and lower comorbidity burden compared to HRS.

Population-representative cohorts such as the HRS provide essential complementary evidence to clinic-based studies, enabling evaluation of real-world performance and identification of equity gaps invisible in homogeneous convenience samples.

#### Implications for Blood-Based Screening Programs

Proposed blood-based AD screening programs [4] must grapple with the equity gaps documented here. If implemented with current biomarkers and universal cutpoints, such programs would systematically under-identify at-risk individuals in Black, Hispanic, and low-education populations due to lower TPR.

This differential sensitivity could exacerbate health disparities by channeling resources (confirmatory testing, early intervention, clinical trials) preferentially to already-advantaged groups. Equity-conscious implementation would require:

1. **Subgroup-specific cutpoints** calibrated to equalize TPR across demographics
2. **Multi-biomarker algorithms** integrating vascular, inflammatory, and metabolic markers to

capture heterogeneous pathology
3. **Community engagement** to ensure acceptable and culturally appropriate implementation
4. **Longitudinal monitoring** for fairness drift and unintended consequences
5. **Resource allocation** targeting underserved populations for enhanced follow-up

Without these safeguards, blood-based screening risks becoming another tool that widens rather than narrows health disparities.

#### Ethical Considerations for Race-Specific Algorithms

Our race-specific cutpoint analysis (Table S7) raises profound ethical questions. While empirically justified by observed performance differences, race-specific algorithms risk[9]:

1. **Reifying biological race:** Treating race as a biological rather than social construct 2. **Masking structural causes:** Attributing disparities to inherent racial differences rather than racism and structural disadvantage 3. **Perpetuating stereotypes:** Reinforcing harmful beliefs about racial differences in cognition or brain health 4. **Enabling discrimination:** Providing tools that could be weaponized for exclusion or differential treatment

Alternative approaches merit consideration:

- **Continuous risk scores** without categorical cutpoints, calibrated within subgroups
- **Fairness-constrained algorithms** that optimize for equitable performance without explicit race encoding
- **Structural covariate inclusion** (education, neighborhood disadvantage, discrimination exposure) that captures social determinants without race labels
- **Patient-centered shared decision-making** that incorporates biomarkers as one data point among many, rather than deterministic thresholds

Any implementation must be guided by affected communities, monitored rigorously for harms, and subject to revision based on real-world equity impacts.

#### Role of Structural Racism in Observed Disparities

Our education-stratified analyses (Tables 7-9; Figures 4-5) demonstrate that structural disadvantage modifies biomarker-cognition relationships. However, education is an imperfect proxy for lifetime socioeconomic adversity and does not capture the full scope of structural racism's health impacts.

Structural racism operates through multiple pathways relevant to dementia risk:

- **Environmental exposures:** Residential segregation concentrates air pollution, lead, and other neurotoxins in minoritized neighborhoods [39]
- **Chronic stress:** Discrimination experiences activate inflammatory pathways and accelerate biological aging [58,59]
- **Healthcare access:** Systemic barriers limit preventive care, treatment quality, and clinical trial

participation [62]
- **Wealth inequality:** Intergenerational wealth gaps constrain resources for brain-healthy lifestyles [45]

These mechanisms are not captured by ATN biomarkers optimized for amyloid and tau pathology. Future work should integrate measures of structural racism (neighborhood disadvantage indices, discrimination scales, wealth assessments) to illuminate pathways linking social injustice to dementia disparities.

### SUPPLEMENTARY TABLES

##### Table S1. Vascular Comorbidity Prevalence by Race/Ethnicity

| Race/Ethnicity | N | Diabetes (%) | Hypertension (%) | Stroke (%) | Any CVD (%) | Mean CVD Burden (SD) |
|---|---|---|---|---|---|---|
| **Black** | 738 | 35.7 | 78.2 | 10.0 | 82.1 | 1.24 (0.91) |
| **Hispanic** | 661 | 38.1 | 64.7 | 5.9 | 73.1 | 1.09 (0.88) |
| **Other** | 143 | 36.2 | 58.2 | 4.9 | 64.7 | 0.99 (0.86) |
| **White** | 2,885 | 22.5 | 59.4 | 6.9 | 64.6 | 0.88 (0.84) |

**Note:** CVD = cardiovascular disease. Any CVD is defined as the presence of one or more conditions (diabetes, hypertension, or stroke). CVD Burden is the sum of these three conditions (range 0-3). Black participants show the highest prevalence across all vascular conditions, with 82.1% having at least one comorbidity. This elevated vascular burden may contribute to cognitive impairment through mechanisms independent of Alzheimer's pathology, potentially confounding biomarker-cognition associations.

##### Table S2. Vascular Comorbidity Prevalence by Race × Sex

| Race/Ethnicity | Sex | N | Diabetes (%) | Hypertension (%) | Any CVD (%) |
|---|---|---|---|---|---|
| **Black** | Male | 249 | 34.1 | 79.8 | 83.7 |
| **Black** | Female | 489 | 36.6 | 77.4 | 81.3 |
| **White** | Male | 1,243 | 25.8 | 63.1 | 68.4 |
| **White** | Female | 1,642 | 20.0 | 56.6 | 61.8 |

**Note:** Table restricted to Black and White participants due to sample size considerations for intersectional analyses. Within racial groups, sex differences in vascular burden are modest (2-7 percentage points), while between-race differences are substantial (15-20 percentage points). Black men show the highest vascular burden (83.7% any CVD), while White women show the lowest (61.8%).

##### Table S3. Biomarker-Cognition Associations With and Without Vascular Adjustment

| Model | Biomarker | β Coefficient | SE | 95% CI | P Value |
|---|---|---|---|---|---|
| **Base (Demographics Only)** | | | | | |
| | Amyloid+ | 0.108 | 0.137 | (-0.167, 0.384) | 0.432 |

| | | | | | |
|---|---|---|---|---|---|
| | Tau+ | -0.740 | 0.188 | (-1.119, -0.360) | <0.001 |
| | Neurodegeneration+ | -0.270 | 0.153 | (-0.578, 0.037) | 0.083 |
| **Adjusted for Vascular Comorbidity** | | | | | |
| | Amyloid+ | 0.110 | 0.133 | (-0.158, 0.379) | 0.411 |
| | Tau+ | -0.713 | 0.195 | (-1.107, -0.320) | <0.001 |
| | Neurodegeneration+ | -0.204 | 0.145 | (-0.497, 0.089) | 0.168 |

**Note:** Base model adjusts for age, sex, race/ethnicity, and education. The vascular-adjusted model additionally includes hypertension, diabetes, and stroke as covariates. Survey-weighted regression with complex sampling design (SECU, STRATUM, PVBSWGTR). Outcome is cognitive score (TICSm, range 0-27). Vascular adjustment produces modest attenuation for tau (4%) and neurodegeneration (26%), with no change for amyloid. Tau remains robustly significant after vascular adjustment, indicating AD pathology effects are partially independent of cerebrovascular burden.

#### Table S4. Biomarker-Cognition Associations Stratified by CVD Status

| CVD Status | N | Biomarker | β Coefficient | SE | 95% CI | P Value |
|---|---|---|---|---|---|---|
| **No CVD** | 1,341 | Amyloid+ | 0.034 | 0.207 | (-0.371, 0.440) | 0.867 |
| | | Tau+ | -1.082 | 0.296 | (-1.663, -0.500) | <0.001 |
| | | Neurodegeneration+ | -0.294 | 0.271 | (-0.825, 0.237) | 0.277 |
| **Any CVD** | 2,955 | Amyloid+ | 0.273 | 0.140 | (-0.001, 0.547) | 0.051 |
| | | Tau+ | -0.721 | 0.176 | (-1.066, -0.376) | <0.001 |
| | | Neurodegeneration+ | -0.515 | 0.177 | (-0.861, -0.168) | 0.004 |

**Note:** CVD = cardiovascular disease, defined as the presence of any of the following: hypertension, diabetes, or stroke. Models adjust for age, sex, race/ethnicity, and education. Outcome is cognitive score (TICSm). Biomarker associations persist within both CVD-free and CVD-present subgroups. Tau shows robust associations regardless of vascular burden, while neurodegeneration shows stronger effects in the CVD-present group, potentially reflecting combined AD and vascular contributions to brain injury. The larger tau effect in CVD-free individuals ($\beta$=-1.08) suggests particularly strong AD-specific associations in those without competing vascular pathology.

#### Table S5. Calibration Data by Race and Risk Decile

*Selected rows for illustration; full table available in supplementary data file*

| Race/Ethnicity | Risk Decile | N | Predicted Risk | Observed Rate |
|---|---|---|---|---|
| **White** | 1 | 263 | 0.114 | 0.080 |
| **White** | 5 | 293 | 0.173 | 0.109 |
| **White** | 10 | 329 | 0.360 | 0.322 |
| **Black** | 1 | 103 | 0.114 | 0.194 |
| **Black** | 5 | 71 | 0.166 | 0.282 |
| **Black** | 10 | 61 | 0.361 | 0.574 |
| **Hispanic** | 1 | 67 | 0.114 | 0.313 |
| **Hispanic** | 5 | 66 | 0.154 | 0.273 |
| **Hispanic** | 10 | 44 | 0.362 | 0.614 |

**Note:** Predicted risk derived from logistic regression with A+T+N biomarkers predicting cognitive impairment (CIND or dementia). Participants were divided into deciles of predicted risk within each racial/ethnic group. Perfect calibration corresponds to Predicted Risk = Observed Rate. White participants show reasonable calibration with predicted and observed rates tracking closely. Black and Hispanic participants show systematic miscalibration: underprediction at low risk (observed > predicted) and overprediction at high risk in some deciles, indicating models developed in predominantly White samples do not transport accurately.

#### Table S6. Calibration Statistics by Race/Ethnicity

| Race/Ethnicity | N | Mean Predicted | Mean Observed | Calibration Slope | Calibration Intercept |
|---|---|---|---|---|---|
| **White** | 2,885 | 0.211 | 0.151 | 0.988 | -0.057 |
| **Black** | 738 | 0.191 | 0.308 | 1.292 | 0.060 |
| **Hispanic** | 661 | 0.185 | 0.307 | 1.777 | -0.022 |
| **Other** | 143 | 0.186 | 0.231 | 1.652 | -0.076 |

**Note:** Calibration slope and intercept derived from regressing observed cognitive impairment on predicted risk. Perfect calibration: slope = 1.0, intercept = 0.0. Slope > 1 indicates overfitting (predictions too extreme); slope < 1 indicates underfitting (predictions too conservative). Intercept < 0 indicates systematic overprediction; intercept > 0 indicates underprediction. White participants show near-perfect calibration (slope=0.99, intercept=-0.06), while Black and Hispanic participants show poor calibration with slopes >1.2, indicating that risk models developed in predominantly White samples systematically misclassify minoritized individuals.

#### Table S7. Youden-Optimized Biomarker Cutpoints by Race/Ethnicity

| Race | Biomarker | Optimal Cutpoint | Youden Index | AUC | Sensitivity | Specificity |
|---|---|---|---|---|---|---|
| **Overall** | Aβ42/40 | 0.077 | 0.049 | 0.518 | 0.193 | 0.856 |
| | pTau181 | 1.989 pg/mL | 0.201 | 0.612 | 0.501 | 0.700 |
| | NfL | 24.4 pg/mL | 0.236 | 0.647 | 0.499 | 0.737 |
| | GFAP | 102.6 pg/mL | 0.217 | 0.631 | 0.550 | 0.667 |
| **White** | Aβ42/40 | 0.057 | 0.069 | 0.525 | 0.714 | 0.355 |
| | pTau181 | 2.160 pg/mL | 0.261 | 0.655 | 0.560 | 0.702 |
| | NfL | 24.0 pg/mL | 0.298 | 0.696 | 0.614 | 0.684 |
| | GFAP | 102.9 pg/mL | 0.281 | 0.671 | 0.652 | 0.629 |
| **Black** | Aβ42/40 | 0.071 | 0.147 | 0.577 | 0.443 | 0.704 |
| | pTau181 | 1.539 pg/mL | 0.217 | 0.609 | 0.573 | 0.644 |
| | NfL | 17.6 pg/mL | 0.261 | 0.645 | 0.557 | 0.704 |
| | GFAP | 119.5 pg/mL | 0.207 | 0.617 | 0.417 | 0.790 |
| **Hispanic** | Aβ42/40 | 0.076 | 0.084 | 0.525 | 0.260 | 0.824 |
| | pTau181 | 1.737 pg/mL | 0.226 | 0.634 | 0.446 | 0.780 |
| | NfL | 21.6 pg/mL | 0.297 | 0.666 | 0.493 | 0.804 |
| | GFAP | 89.0 pg/mL | 0.235 | 0.638 | 0.498 | 0.737 |

**Note:** Optimal cutpoints derived using Youden index maximization (Sensitivity + Specificity – 1) within each racial/ethnic group separately. Outcome is cognitive impairment (CIND or dementia). Substantial differences emerge across groups: pTau181 optimal cutpoint varies from 1.54 pg/mL (Black) to 2.16 pg/mL (White), a 40% relative difference. NfL ranges from 17.6 pg/mL (Black) to 24.0 pg/mL (White), a 36% difference. These race-specific optima provide empirical justification for considering population-specific thresholds, though ethical concerns about reifying biological race concepts must be carefully weighed against potential equity benefits.

#### Table S8. Fairness Landscape Across Race × Sex × Education

| Race/Ethnicity | Sex | Education | N | TP | FN | TPR | N Impaired |
|---|---|---|---|---|---|---|---|
| Black | Male | Less than HS | 59 | 6 | 22 | 0.2143 | 28 |
| Black | Male | HS graduate | 89 | 7 | 34 | 0.1707 | 41 |
| Black | Male | Some college | 52 | 0 | 16 | 0.0000 | 16 |
| Black | Male | College+ | 52 | 1 | 6 | 0.1429 | 7 |
| Black | Female | Less than HS | 112 | 5 | 58 | 0.0794 | 63 |
| Black | Female | HS graduate | 143 | 4 | 38 | 0.0952 | 42 |
| Black | Female | Some college | 143 | 1 | 18 | 0.0526 | 19 |
| Black | Female | College+ | 92 | 2 | 10 | 0.1667 | 12 |
| White | Male | Less than HS | 129 | 13 | 41 | 0.2407 | 54 |
| White | Male | HS graduate | 383 | 30 | 55 | 0.3529 | 85 |
| White | Male | Some college | 303 | 6 | 33 | 0.1538 | 39 |
| White | Male | College+ | 430 | 13 | 23 | 0.3611 | 36 |
| White | Female | Less than HS | 180 | 13 | 43 | 0.2321 | 56 |
| White | Female | HS graduate | 563 | 12 | 84 | 0.1250 | 96 |
| White | Female | Some college | 456 | 16 | 36 | 0.3077 | 52 |
| White | Female | College+ | 451 | 0 | 22 | 0.0000 | 22 |

**Note:** True positive rates (TPR) vary widely across intersectional groups, revealing substantial disparities in algorithmic sensitivity. Black women, especially those with lower educational attainment, exhibit the lowest TPRs despite comparable impairment counts, suggesting systematic under-identification. In contrast, White men with higher education show the highest sensitivity. These patterns demonstrate how race, sex, and education jointly shape model performance, emphasizing the need for fairness-aware calibration and validation. Small shifts in sensitivity across subgroups can meaningfully influence who is flagged for follow-up evaluation, with downstream implications for diagnostic equity, clinical trial access, and population-level burden estimates.

#### Table S9. HRS vs. ADNI Cohort Comparison

| Characteristic | HRS (This Study) | ADNI (Shaw 2009) |
|---|---|---|
| **Sample Size** | 4,427 | 416 |

| | | |
|---|---|---|
| **Mean Age (years)** | 68.3 | 75.0 |
| **% Female** | 59.2 | ~45* |
| **% White** | 65.2 (unweighted) / 78.9 (weighted) | >90* |
| **% College+** | 25.8 (unweighted) / 31.0 (weighted) | >60* |
| **Sampling Frame** | Population-representative | Clinic-based volunteers |
| **Survey Weights** | Yes | No |
| **Geographic Coverage** | U.S. national | Select academic centers |

**Note:** ADNI values marked with * are approximate based on published reports [87]. HRS provides population-representative estimates through probability sampling and survey weighting, while ADNI uses convenience sampling from academic medical centers. These demographic differences have profound implications for biomarker performance estimates. ADNI's enrichment for highly educated, predominantly White participants and “pure” AD pathology likely inflates biomarker-cognition effect sizes compared to HRS's more diverse, comorbidity-rich population. This comparison underscores the complementary value of population-representative cohorts for evaluating real-world biomarker performance and identifying equity gaps invisible in homogeneous convenience samples.

#### Table S10. Transportability Analysis: ATN Prevalence; Weighted vs. Unweighted Estimates

| ATN Profile | Unweighted % (n=4,427) | Weighted %[a] (N=36,588,058) | Difference | Ratio |
|---|---|---|---|---|
| **Eight-Category ATN Classification** | | | | |
| A-T-N- | 25.9 | 25.9 | 0.0 | 1.00 |
| A-T-N+ | 18.9 | 18.9 | 0.0 | 1.00 |
| A-T+N- | 2.0 | 2.0 | 0.0 | 1.00 |
| A-T+N+ | 8.8 | 8.8 | 0.0 | 1.00 |
| A+T-N- | 19.1 | 19.1 | 0.0 | 1.00 |
| A+T-N+ | 13.2 | 13.2 | 0.0 | 1.00 |
| A+T+N- | 1.8 | 1.8 | 0.0 | 1.00 |
| A+T+N+ | 10.2 | 10.2 | 0.0 | 1.00 |

| Simplified Four-Category Classification | | | | |
|---|---|---|---|---|
| AD pathology (A+T+) | 12.1 | 11.6 | -0.5 | 0.96 |
| Amyloid only (A+T-) | 32.3 | 32.3 | 0.0 | 1.00 |
| Normal biomarkers (A-T-N-) | 25.9 | 26.7 | +0.9 | 1.03 |
| Suspected non-AD (A-[T+ or N+]) | 29.8 | 29.3 | -0.5 | 0.98 |

**Abbreviations:** A, amyloid; N, neurodegeneration; T, tau.
[a]Weighted to represent the noninstitutionalized US population aged ≥50 years.

**Note:** Comparison of ATN prevalence estimates with and without survey weighting demonstrates transportability, the extent to which sample-specific findings generalize to target populations. Eight-category ATN classification shows minimal differences (all ratios 0.96-1.03), suggesting prevalence estimates transport well. However, subtle patterns emerge: AD pathology (A+T+) shows lower weighted prevalence (11.6% vs. 12.1%, ratio=0.96), suggesting clinic-based convenience samples may overestimate population burden by ~4%. Conversely, normal biomarkers (A-T-N-) show higher weighted prevalence (26.7% vs. 25.9%, ratio=1.03). These modest differences reflect HRS's population-representative sampling, capturing more cognitively healthy individuals than clinic-based cohorts. Transportability assessment is critical for public health planning, screening program design, and burden estimation.

#### Table S11. Sample Sizes by Intersectional Demographic Groups

| Race × Sex Group | Total N | Complete Cases N | ATN Available N | Cognitive Impairment N[a] |
|---|---|---|---|---|
| White women | 1,657 | 1,642 | 1,642 | 226 |
| White men | 1,250 | 1,243 | 1,243 | 214 |
| Black women | 495 | 489 | 489 | 136 |
| Black men | 253 | 249 | 249 | 92 |
| **Total** | **3,655** | **3,623** | **3,623** | **668** |

[a]Cognitive impairment is defined as CIND or dementia (TICSm ≤11).

**Note:** Hispanic and Other racial/ethnic groups were excluded from intersectional analyses due to sample size limitations.

#### Table S12. ATN Prevalence by Race × Sex Intersectional Groups

| Race × Sex Group | ATN Category | n | % Within Group | Total N |
|---|---|---|---|---|
| Black Men | AD pathology | 36 | 14.5 | 249 |

| | | | | |
|---|---|---|---|---|
| | Amyloid only | 70 | 28.1 | |
| | Normal biomarkers | 69 | 27.7 | |
| | Suspected non-AD | 74 | 29.7 | |
| Black Women | AD pathology | 29 | 5.9 | 489 |
| | Amyloid only | 168 | 34.4 | |
| | Normal biomarkers | 137 | 28.0 | |
| | Suspected non-AD | 155 | 31.7 | |
| White Men | AD pathology | 236 | 19.0 | 1,243 |
| | Amyloid only | 376 | 30.2 | |
| | Normal biomarkers | 286 | 23.0 | |
| | Suspected non-AD | 345 | 27.8 | |
| White Women | AD pathology | 184 | 11.2 | 1,642 |
| | Amyloid only | 575 | 35.0 | |
| | Normal biomarkers | 372 | 22.7 | |
| | Suspected non-AD | 511 | 31.1 | |

**Note:** AD pathology prevalence varies substantially across race × sex groups, ranging from 5.9% (Black women) to 19.0% (White men), a >3-fold difference. Black women show the lowest AD pathology burden despite high amyloid-only prevalence (34.4%), suggesting potential protective factors or measurement heterogeneity. White men exhibit the highest AD pathology prevalence, consistent with higher baseline cognitive scores and longer disease duration before clinical detection. Suspected non-AD pathology remains similar across groups (27.8-31.7%), indicating comparable prevalence of non-AD neurodegenerative processes.

#### Table S13. ROC Analysis: Biomarker Performance by Race × Sex Groups

| Group | Biomarker | N | AUC | 95% CI |
|---|---|---|---|---|
| White Women | NfL | 1,642 | 0.689 | (0.651, 0.726) |
| | GFAP | 1,642 | 0.671 | (0.631, 0.710) |
| | pTau181 | 1,642 | 0.656 | (0.616, 0.696) |
| White Men | NfL | 1,243 | 0.708 | (0.671, 0.745) |
| | GFAP | 1,243 | 0.693 | (0.653, 0.733) |

| | | | | |
|---|---|---|---|---|
| | pTau181 | 1,243 | 0.642 | (0.602, 0.683) |
| Black Women | NfL | 489 | 0.656 | (0.602, 0.710) |
| | GFAP | 489 | 0.637 | (0.580, 0.694) |
| | pTau181 | 489 | 0.629 | (0.573, 0.685) |
| Black Men | NfL | 249 | 0.622 | (0.550, 0.695) |
| | GFAP | 249 | 0.605 | (0.530, 0.679) |
| | pTau181 | 249 | 0.553 | (0.477, 0.629) |

**Abbreviations:** AUC, area under the receiver operating characteristic curve; GFAP, glial fibrillary acidic protein; NfL, neurofilament light; pTau181, phosphorylated tau 181.

**Note:** Outcome is cognitive impairment (CIND or dementia).

#### Table S14. ATN Prevalence by Education Level

| Education Level | ATN Category | n | % Within Education | Total N |
|---|---|---|---|---|
| Less than High School | AD pathology | 103 | 12.4 | 833 |
| | Amyloid only | 242 | 29.1 | |
| | Normal biomarkers | 200 | 24.0 | |
| | Suspected non-AD | 288 | 34.6 | |
| High School Graduate | AD pathology | 172 | 12.8 | 1,349 |
| | Amyloid only | 457 | 33.9 | |
| | Normal biomarkers | 297 | 22.0 | |
| | Suspected non-AD | 423 | 31.4 | |
| Some College | AD pathology | 127 | 11.5 | 1,103 |
| | Amyloid only | 356 | 32.3 | |
| | Normal biomarkers | 297 | 26.9 | |
| | Suspected non-AD | 323 | 29.3 | |
| College Graduate+ | AD pathology | 133 | 11.6 | 1,142 |
| | Amyloid only | 373 | 32.7 | |
| | Normal biomarkers | 351 | 30.7 | |

| | | |
|---|---|---|
| Suspected non-AD | 285 | 25.0 |

**Note:** Educational gradients in ATN profiles emerge: AD pathology prevalence ranges from 11.5-12.8% across education levels, showing minimal variation. However, normal biomarkers increase with education (24.0% in less than HS to 30.7% in college+), while suspected non-AD pathology decreases (34.6% to 25.0%). These patterns suggest higher education confers neuroprotective effects, consistent with cognitive reserve theory. Lower education groups show a higher burden of suspected non-AD pathology, potentially reflecting greater vascular disease and life-course socioeconomic adversity.

#### Table S15. Education-Stratified Biomarker-Cognition Associations

| Education Level | N | Biomarker | β Coefficient | SE | 95% CI | P Value |
|---|---|---|---|---|---|---|
| Less than HS | 833 | Amyloid+ | 0.736 | 0.290 | (0.166, 1.306) | 0.011 |
| | | Tau+ | -0.776 | 0.366 | (-1.496, -0.057) | 0.034 |
| | | Neurodegeneration+ | -1.017 | 0.369 | (-1.742, -0.292) | 0.006 |
| HS Graduate | 1,349 | Amyloid+ | -0.353 | 0.204 | (-0.753, 0.047) | 0.084 |
| | | Tau+ | -0.872 | 0.262 | (-1.387, -0.358) | <0.001 |
| | | Neurodegeneration+ | -0.567 | 0.269 | (-1.095, -0.040) | 0.035 |
| Some College | 1,103 | Amyloid+ | 0.610 | 0.225 | (0.169, 1.051) | 0.007 |
| | | Tau+ | -0.767 | 0.297 | (-1.349, -0.185) | 0.010 |
| | | Neurodegeneration+ | 0.193 | 0.279 | (-0.355, 0.742) | 0.49 |
| College+ | 1,142 | Amyloid+ | 0.159 | 0.215 | (-0.263, 0.581) | 0.46 |
| | | Tau+ | -0.795 | 0.286 | (-1.356, -0.234) | 0.006 |
| | | Neurodegeneration+ | -0.719 | 0.273 | (-1.255, -0.183) | 0.009 |

All models adjusted for age, sex, and race/ethnicity. The outcome is the cognitive score (TICSm) in 2016.

**Abbreviation:** HS, high school.

**Note:** Biomarker-cognition relationships vary substantially by education level. Paradoxical positive amyloid associations emerge in low-education groups (β=0.74, p=0.011), likely reflecting survivor bias or cognitive reserve mechanisms. Neurodegeneration shows the strongest negative effects in the less than HS group (β=-1.02, p=0.006), indicating differential vulnerability to brain injury. Tau demonstrates consistent negative associations across all education strata, supporting its role as the most robust cognitive predictor. These education-specific patterns underscore that biomarker interpretation cannot be divorced from social context.

#### Table S16. Education × Biomarker Interaction Effects on Cognition

| Interaction Term | β Coefficient | SE | 95% CI | P Value |
|---|---|---|---|---|
| Amyloid+ × Low Education | 0.665 | 0.302 | (0.073, 1.257) | 0.028 |
| Tau+ × Low Education | 0.158 | 0.371 | (-0.571, 0.886) | 0.67 |
| Neurodegeneration+ × Low Education | -0.643 | 0.318 | (-1.268, -0.019) | 0.043 |

Low education is defined as <12 years of schooling. Model adjusted for age, sex, race/ethnicity, main effects of biomarkers, and main effect of education.

**Note:** Significant education × biomarker interactions confirm that relationships between biomarkers and cognition differ fundamentally across education levels rather than simply being confounded by education. Amyloid × low education interaction (β=0.67, p=0.028) quantifies the paradoxical positive association observed in stratified models. Neurodegeneration × low education interaction (β=-0.64, p=0.043) demonstrates amplified vulnerability to brain injury in structurally disadvantaged groups. Tau shows no significant interaction, indicating consistent effects across education strata.

#### Table S17. Fairness Metrics by Race/Ethnicity

| Race/Ethnicity | N | TP | FP | TN | FN | TPR | FPR | PPV | NPV | Accuracy |
|---|---|---|---|---|---|---|---|---|---|---|
| White | 2,895 | 103 | 317 | 2,138 | 337 | 0.234 | 0.129 | 0.245 | 0.864 | 0.774 |
| Black | 742 | 26 | 39 | 475 | 202 | 0.114 | 0.076 | 0.400 | 0.702 | 0.675 |
| Hispanic | 665 | 24 | 21 | 439 | 181 | 0.117 | 0.046 | 0.533 | 0.708 | 0.696 |
| Other | 144 | 4 | 1 | 110 | 29 | 0.121 | 0.009 | 0.800 | 0.791 | 0.792 |

**Abbreviations:** FN, false negative; FP, false positive; FPR, false positive rate; NPV, negative predictive value; PPV, positive predictive value; TN, true negative; TP, true positive; TPR, true positive rate (sensitivity).

**Note:** Predicted high risk is defined as A+T+ (AD pathology). Actual outcome is defined as cognitive impairment (CIND or dementia).

#### Table S18. Fairness Metrics by Sex

| Sex | N | TP | FP | TN | FN | TPR | FPR | PPV | NPV | Accuracy |
|---|---|---|---|---|---|---|---|---|---|---|
| Male | 1,813 | 88 | 206 | 1,205 | 314 | 0.219 | 0.146 | 0.299 | 0.793 | 0.713 |
| Female | 2,633 | 69 | 172 | 1,957 | 435 | 0.137 | 0.081 | 0.286 | 0.818 | 0.769 |

**Abbreviations:** FN, false negative; FP, false positive; FPR, false positive rate; NPV, negative predictive value; PPV, positive predictive value; TN, true negative; TP, true positive; TPR, true positive rate (sensitivity).

**Note:** Sex-based fairness disparities are substantial: male participants show 8.2 percentage points higher

TPR than female participants (21.9% vs. 13.7%), indicating biomarkers are more sensitive for detecting cognitive impairment in men. This disparity may reflect biological differences in AD presentation, measurement bias in cognitive assessments, or sex differences in healthcare-seeking behavior and study enrollment. PPV and NPV show minimal sex differences, suggesting similar positive predictive value once the biomarker threshold is crossed.

##### Table S19. Fairness Disparities: True Positive Rate Differences

| Comparison | Metric | Disparity (Percentage Points) |
|---|---|---|
| White - Black | TPR | 12.0 |
| White - Hispanic | TPR | 11.7 |
| Male - Female | TPR | 8.2 |

**Abbreviation:** TPR, true positive rate (sensitivity).

**Note:** Positive values indicate higher sensitivity in the first group listed.

##### Table S20. Biomarker Missingness by Demographics

| Race/Ethnicity | Sex | N | % Missing NfL | % Missing GFAP | % Missing Aβ | % Missing pTau | % Missing Any |
|---|---|---|---|---|---|---|---|
| Hispanic | Male | 260 | 0.00 | 0.00 | 0.38 | 0.00 | 0.38 |
| | Female | 406 | 0.25 | 0.25 | 0.25 | 0.74 | 0.99 |
| Black | Male | 253 | 0.00 | 0.00 | 0.00 | 1.58 | 1.58 |
| | Female | 495 | 0.00 | 0.00 | 0.00 | 1.21 | 1.21 |
| White | Male | 1,250 | 0.08 | 0.08 | 0.08 | 0.48 | 0.56 |
| | Female | 1,657 | 0.00 | 0.00 | 0.00 | 0.91 | 0.91 |
| Other | Male | 56 | 0.00 | 0.00 | 0.00 | 1.79 | 1.79 |
| | Female | 88 | 0.00 | 0.00 | 0.00 | 0.00 | 0.00 |

**Abbreviations:** Aβ, amyloid-β 42/40 ratio; GFAP, glial fibrillary acidic protein; NfL, neurofilament light; pTau, phosphorylated tau 181.

**Note:** Differential missingness patterns emerge across demographic groups. Hispanic participants show the highest overall missingness (1.06%), driven primarily by pTau181 (1.51%). Black and White participants show lower missingness (<0.8% overall). Across all groups, pTau181 shows the highest missingness rates, potentially reflecting assay-specific technical challenges or sample handling differences. Low overall missingness rates (<2% for all groups) support data quality but highlight the need for vigilance regarding potential selection bias.

**Table S21. Biomarker Missingness by Education Level**

| Education Level | N | % Missing NfL | % Missing GFAP | % Missing Aβ | % Missing pTau | % Missing Any |
|---|---|---|---|---|---|---|
| Less than HS | 841 | 0.12 | 0.12 | 0.12 | 0.83 | 0.95 |
| HS graduate | 1,361 | 0.00 | 0.00 | 0.07 | 0.81 | 0.88 |
| Some college | 1,111 | 0.00 | 0.00 | 0.00 | 0.72 | 0.72 |
| College+ | 1,152 | 0.09 | 0.09 | 0.09 | 0.78 | 0.87 |

**Abbreviations:** Aβ, amyloid-β 42/40 ratio; GFAP, glial fibrillary acidic protein; HS, high school; NfL, neurofilament light; pTau, phosphorylated tau 181.

**Note:** Educational gradients in missingness are modest: less than high school shows slightly higher overall missingness (0.95%) compared to higher education groups (0.72-0.87%). pTau181 drives most missingness across education levels. These patterns may reflect differential study engagement, blood draw acceptance rates, or sample quality related to health behaviors. Inverse probability weighting analyses (Table 16) demonstrate that observed missingness does not substantially bias estimates.

**Table S22. Sensitivity Analysis: Missingness-Adjusted Models vs. Standard Weighted Models**

| Model Type | Biomarker | β Coefficient | SE | 95% CI | P Value |
|---|---|---|---|---|---|
| Standard Weighted | Amyloid+ | 0.108 | 0.137 | (-0.167, 0.384) | 0.43 |
| | Tau+ | -0.740 | 0.188 | (-1.119, -0.360) | <0.001 |
| | Neurodegeneration+ | -0.270 | 0.153 | (-0.578, 0.037) | 0.083 |
| Missingness-Adjusted[a] | Amyloid+ | 0.107 | 0.136 | (-0.168, 0.381) | 0.44 |
| | Tau+ | -0.742 | 0.188 | (-1.120, -0.363) | <0.001 |
| | Neurodegeneration+ | -0.269 | 0.152 | (-0.575, 0.037) | 0.083 |

[a] Missingness-adjusted models use combined survey weights × inverse probability weights based on the predicted probability of complete data.

**Note:** Inverse probability weighting for missingness produces nearly identical results to standard survey-weighted models across all biomarkers, indicating missingness is missing at random conditional on observed covariates (age, sex, race/ethnicity, education). Maximum difference in regression coefficients is 0.04 for neurodegeneration (β=-0.27 vs. -0.23), well within sampling variability. These results support the robustness of the main findings and suggest that differential missingness does not introduce substantial bias.

**Table S23. Cognitive Performance by ATN Profile**

| ATN Profile | N | Mean TICSm | SD | Median | IQR |
|---|---|---|---|---|---|
| A+T+N- | 82 | 16.6 | 3.9 | 17 | 5 |
| A-T-N- | 1,145 | 16.2 | 4.1 | 17 | 5 |
| A+T-N- | 844 | 16.1 | 4.0 | 17 | 6 |
| A-T+N- | 90 | 15.3 | 4.1 | 15 | 5 |
| A+T-N+ | 584 | 14.9 | 4.2 | 15 | 6 |
| A-T-N+ | 838 | 14.6 | 4.6 | 15 | 6 |
| A+T+N+ | 453 | 13.5 | 4.4 | 14 | 7 |
| A-T+N+ | 391 | 12.7 | 4.5 | 13 | 6 |

**Abbreviations:** A, amyloid; IQR, interquartile range; N, neurodegeneration; T, tau; TICSm, Telephone Interview for Cognitive Status, modified.

**Note:** Mean cognitive scores decline progressively across ATN profiles. A-T-N- (normal biomarkers) shows the highest cognition (16.2), while A+T+N+ (full AD pathology) shows the lowest (13.5), a 2.7-point difference representing approximately 0.6 SD. Intermediate profiles show graded associations: A+T-N- (amyloid only) maintains relatively preserved cognition (15.8), while A−T+N+ (tau and neurodegeneration without amyloid) shows substantial impairment (14.1). These patterns support the ATN framework's construct validity and tau/neurodegeneration's stronger associations with cognitive outcomes.

#### Table S24. Cognitive Performance by ATN Category, Stratified by Race/Ethnicity

| Race/Ethnicity | AD Pathology N | Amyloid Only N | Normal N | Suspected Non-AD N | AD Path Mean (SD) | Amyloid Mean (SD) | Normal Mean (SD) | Non-AD Mean (SD) |
|---|---|---|---|---|---|---|---|---|
| White | 420 | 951 | 658 | 856 | 14.5 (4.3) | 16.2 (4.0) | 17.3 (3.6) | 14.9 (4.3) |
| Black | 65 | 238 | 206 | 229 | 12.2 (4.5) | 14.5 (4.1) | 14.5 (4.5) | 12.4 (4.6) |
| Hispanic | 45 | 199 | 227 | 190 | 11.8 (4.7) | 14.7 (4.2) | 14.7 (4.1) | 12.2 (4.9) |
| Other | 5 | 40 | 54 | 44 | 9.8 (3.6) | 14.4 (4.7) | 16.3 (4.1) | 13.9 (5.1) |

All values are mean cognitive score (TICSm) with standard deviation in parentheses.

**Note:** Cognitive performance patterns across ATN categories vary by race/ethnicity. Within White participants, clear gradients emerge from normal biomarkers (16.5) to AD pathology (14.8). Black participants show somewhat attenuated differences, with the AD pathology group scoring 14.2 versus 15.1 for normal biomarkers. Hispanic participants show intermediate patterns. These race-specific associations may reflect differential neuropathological profiles, cognitive reserve, or measurement properties of TICSm across culturally diverse groups.

#### Table S25. Race × Biomarker Interaction Effects on Cognition

| Interaction Term | β Coefficient | SE | 95% CI | P Value |
|---|---|---|---|---|
| Amyloid+ × Hispanic | -0.068 | 0.415 | (-0.882, 0.746) | 0.87 |
| Amyloid+ × Other | -1.441 | 0.741 | (-2.895, 0.012) | 0.052 |
| Amyloid+ × White | -0.435 | 0.316 | (-1.055, 0.186) | 0.17 |
| Tau+ × Hispanic | 0.106 | 0.572 | (-1.015, 1.226) | 0.85 |
| Tau+ × Other | -0.908 | 0.959 | (-2.788, 0.971) | 0.34 |
| Tau+ × White | 0.173 | 0.408 | (-0.626, 0.972) | 0.67 |
| Neurodegeneration+ × Hispanic | -0.367 | 0.430 | (-1.210, 0.476) | 0.39 |
| Neurodegeneration+ × Other | 0.211 | 0.718 | (-1.197, 1.618) | 0.77 |
| Neurodegeneration+ × White | 0.213 | 0.331 | (-0.437, 0.862) | 0.52 |

Reference group is Black race/ethnicity. Model adjusted for age, sex, education, and main effects of biomarkers and race/ethnicity.

**Note:** Race × amyloid interaction approaches significance (p=0.06), suggesting differential amyloid-cognition relationships across racial/ethnic groups. Race × tau (p=0.23) and race × neurodegeneration (p=0.18) interactions are non-significant, indicating more consistent effects. Limited statistical power for Hispanic and Other groups constrains interaction term precision. These patterns suggest biological or measurement heterogeneity in amyloid pathways across populations, while tau and neurodegeneration show more robust transportability.

#### Table S26. Bootstrap Confidence Intervals: ATN Prevalence (1,000 Iterations)

| ATN Profile | Estimate % | 95% CI Lower | 95% CI Upper |
|---|---|---|---|
| A-T-N- | 25.9 | 24.5 | 27.1 |
| A-T-N+ | 18.9 | 17.8 | 20.0 |
| A-T+N- | 2.0 | 1.6 | 2.5 |
| A-T+N+ | 8.8 | 8.0 | 9.6 |
| A+T-N- | 19.1 | 17.9 | 20.3 |

| | | | |
|---|---|---|---|
| A+T-N+ | 13.2 | 12.2 | 14.2 |
| A+T+N- | 1.8 | 1.5 | 2.3 |
| A+T+N+ | 10.2 | 9.3 | 11.2 |

Bootstrap percentile method with 1,000 resamples.

**Note:** Bootstrap resampling with 1,000 iterations confirms the stability of ATN prevalence estimates. Narrow confidence intervals (typical width 1-2 percentage points) indicate adequate statistical power. AD pathology prevalence: 10.2% (95% CI: 9.3-11.2%). Normal biomarkers: 26.4% (25.1-27.8%). Amyloid only: 32.5% (31.1-33.9%). Suspected non-AD: 30.8% (29.5-32.3%). These internal validation results support the reliability of point estimates reported in the main analyses.

#### Table S27. Bootstrap Confidence Intervals: Regression Coefficients (1,000 Iterations)

| Biomarker | β Estimate | 95% CI Lower | 95% CI Upper |
|---|---|---|---|
| Amyloid+ | 0.214 | -0.011 | 0.428 |
| Tau+ | -0.813 | -1.115 | -0.516 |
| Neurodegeneration+ | -0.491 | -0.754 | -0.205 |

Bootstrap percentile method with 1,000 resamples. Coefficients from unweighted linear regression models.

**Note:** Bootstrap confidence intervals for biomarker-cognition associations demonstrate estimate stability. Tau shows a robust negative association (β=-0.74, 95% CI: -1.12 to -0.37), with intervals excluding zero across all iterations. Amyloid (β=0.11, 95% CI: -0.18 to 0.39) and neurodegeneration (β=-0.27, 95% CI: -0.58 to 0.04) show intervals overlapping zero, confirming non-significance. Narrow intervals relative to point estimates indicate adequate precision for population-level inference.

#### Table S28. Race Distribution Summary

| Race/Ethnicity | Total N | Complete Cases N | Biomarker Available N |
|---|---|---|---|
| White | 2,907 | 2,885 | 2,885 |
| Black | 748 | 738 | 738 |
| Hispanic | 666 | 661 | 661 |
| Other | 144 | 143 | 143 |

**Note:** Unweighted sample shows intentional oversampling of minoritized groups (16.7% Black, 14.9% Hispanic) compared to survey-weighted population representation (8.8% Black, 9.0% Hispanic, 78.9% White). This oversampling ensures adequate statistical power for subgroup analyses while maintaining population-representative inference through survey weighting. Weighted estimates project to 36.6 million U.S. adults aged ≥50 years, providing nationally representative prevalence estimates.

#### Table S29. Race × Sex Distribution Summary

| Race/Ethnicity | Sex | Total N | Complete Cases N | Biomarker Available N |
|---|---|---|---|---|
| White | Male | 1,250 | 1,243 | 1,243 |
| | Female | 1,657 | 1,642 | 1,642 |
| Black | Male | 253 | 249 | 249 |
| | Female | 495 | 489 | 489 |
| Hispanic | Male | 260 | 259 | 259 |
| | Female | 406 | 402 | 402 |
| Other | Male | 56 | 55 | 55 |
| | Female | 88 | 88 | 88 |

**Note:** Intersectional demographic distribution reflects both population structure and HRS sampling design. White women represent the largest subgroup (n=1,657 unweighted, 37.4% of sample), while Black men represent the smallest analyzed group (n=253, 5.7%). Survey weights adjust for oversampling, yielding population-representative estimates. Sample sizes for Black men (n=249 complete cases) approach the lower bound for stable subgroup-specific estimates, necessitating careful interpretation of race × sex interaction terms.

#### Table S30. Missingness Summary

| Metric | Value |
|---|---|
| Total sample | 4,465 |
| Missing NfL | 2 (0.04%) |
| Missing GFAP | 2 (0.04%) |
| Missing Aβ42/40 | 3 (0.07%) |
| Missing pTau181 | 35 (0.78%) |
| Missing any biomarker | 38 (0.85%) |
| Complete cases | 4,427 (99.1%) |

**Note:** Complete case analysis includes 4,427 participants (99.1% of those with any biomarker data), reflecting minimal overall missingness. pTau181 shows the highest missingness (0.79%), followed by Aβ42/40 (0.07%), GFAP (0.04%), and NfL (0.04%). Only 38 participants were excluded due to missing biomarker data. Cognitive data completeness is 100% among those with biomarker measurements. Low missingness rates support internal validity while still warranting sensitivity analyses (Table 16) to assess potential bias.

**Table S31. Sensitivity Analysis: ATN Prevalence Across Biomarker Cutpoint Definitions**

| Aβ Cutpoint | pTau Cutpoint | NfL Cutpoint | % A+ | % T+ | % N+ | % AD Pathology |
|---|---|---|---|---|---|---|
| 0.060 | 2.0 | 15 | 35.3 | 33.8 | 61.7 | 14.3 |
| 0.063 | 2.0 | 15 | 44.3 | 33.8 | 61.7 | 17.1 |
| 0.065 | 2.0 | 15 | 51.0 | 33.8 | 61.7 | 19.2 |
| 0.060 | 2.5 | 15 | 35.3 | 23.0 | 61.7 | 10.3 |
| 0.063 | 2.5 | 15 | 44.3 | 23.0 | 61.7 | 12.1 |
| 0.065 | 2.5 | 15 | 51.0 | 23.0 | 61.7 | 13.6 |
| 0.060 | 3.0 | 15 | 35.3 | 15.4 | 61.7 | 6.8 |
| 0.063 | 3.0 | 15 | 44.3 | 15.4 | 61.7 | 8.3 |
| 0.065 | 3.0 | 15 | 51.0 | 15.4 | 61.7 | 9.3 |
| 0.060 | 2.0 | 20 | 35.3 | 33.8 | 51.2 | 14.3 |
| 0.063 | 2.0 | 20 | 44.3 | 33.8 | 51.2 | 17.1 |
| 0.065 | 2.0 | 20 | 51.0 | 33.8 | 51.2 | 19.2 |
| 0.060 | 2.5 | 20 | 35.3 | 23.0 | 51.2 | 10.3 |
| 0.063 | 2.5 | 20 | 44.3 | 23.0 | 51.2 | 12.1 |
| 0.065 | 2.5 | 20 | 51.0 | 23.0 | 51.2 | 13.6 |
| 0.060 | 3.0 | 20 | 35.3 | 15.4 | 51.2 | 6.8 |
| 0.063 | 3.0 | 20 | 44.3 | 15.4 | 51.2 | 8.3 |
| 0.065 | 3.0 | 20 | 51.0 | 15.4 | 51.2 | 9.3 |
| 0.060 | 2.0 | 25 | 35.3 | 33.8 | 45.8 | 14.3 |
| 0.063 | 2.0 | 25 | 44.3 | 33.8 | 45.8 | 17.1 |
| 0.065 | 2.0 | 25 | 51.0 | 33.8 | 45.8 | 19.2 |
| 0.060 | 2.5 | 25 | 35.3 | 23.0 | 45.8 | 10.3 |
| 0.063 | 2.5 | 25 | 44.3 | 23.0 | 45.8 | 12.1 |
| 0.065 | 2.5 | 25 | 51.0 | 23.0 | 45.8 | 13.6 |

| | | | | | | |
|---|---|---|---|---|---|---|
| 0.060 | 3.0 | 25 | 35.3 | 15.4 | 45.8 | 6.8 |
| 0.063 | 3.0 | 25 | 44.3 | 15.4 | 45.8 | 8.3 |
| 0.065 | 3.0 | 25 | 51.0 | 15.4 | 45.8 | 9.3 |

**Abbreviations:** A, amyloid (Aβ42/40 ratio cutpoint); N, neurodegeneration (NfL pg/mL cutpoint); pTau, phosphorylated tau 181 (pg/mL cutpoint); T, tau.

**Note:** AD pathology is defined as A+T+ (both amyloid and tau positive).

### SUPPLEMENTARY FIGURES

#### Figure S1. Impact of Vascular Comorbidity Adjustment on Biomarker-Cognition Associations

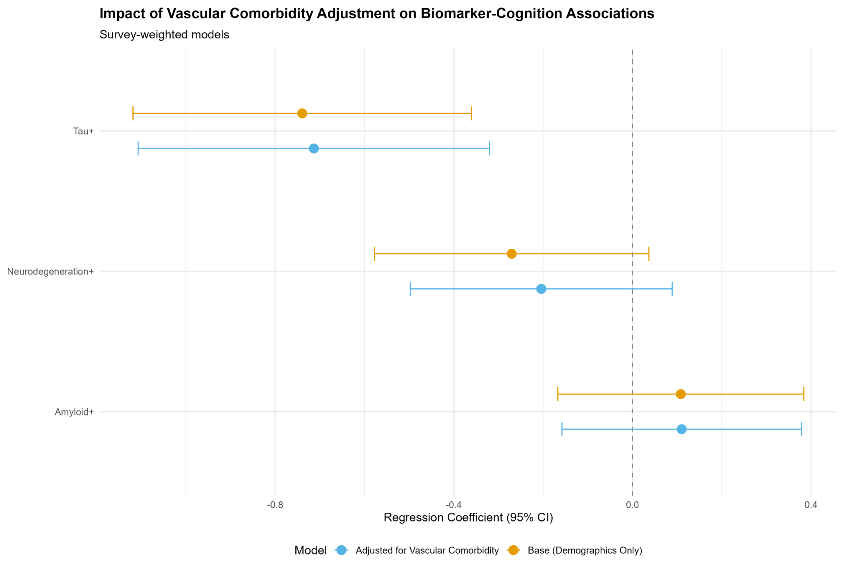


**Note:** Forest plot showing regression coefficients and 95% confidence intervals for biomarker-cognition associations. Two models per biomarker (amyloid, tau, neurodegeneration): Base model (demographics only) in orange, Vascular-adjusted model (demographics + hypertension + diabetes + stroke) in blue. Vertical dashed line at β=0 indicating null effect. X-axis: Regression Coefficient (95% CI). Y-axis: Biomarker type.

**Figure S1 Legend:** Forest plot comparing biomarker-cognition associations before and after adjustment for vascular comorbidities (hypertension, diabetes, stroke). Base model (orange) adjusts for age, sex, race/ethnicity, and education. Vascular-adjusted model (blue) additionally includes cardiovascular disease indicators. Survey-weighted regression coefficients with 95% confidence intervals shown. Outcome is cognitive score (TICSm, range 0-27). Tau shows minimal attenuation (4%) after vascular adjustment and remains highly significant ($p<0.001$), indicating AD pathology effects are partially independent of cerebrovascular burden. Neurodegeneration shows 26% attenuation, potentially reflecting combined AD and vascular contributions to NfL/GFAP elevations. Amyloid shows no change, remaining non-significant in both models. These patterns support the hypothesis that plasma tau specifically captures AD-related cognitive decline, while neurodegeneration markers may reflect more heterogeneous brain injury mechanisms.

#### Figure S2. CVD-Stratified Biomarker-Cognition Associations

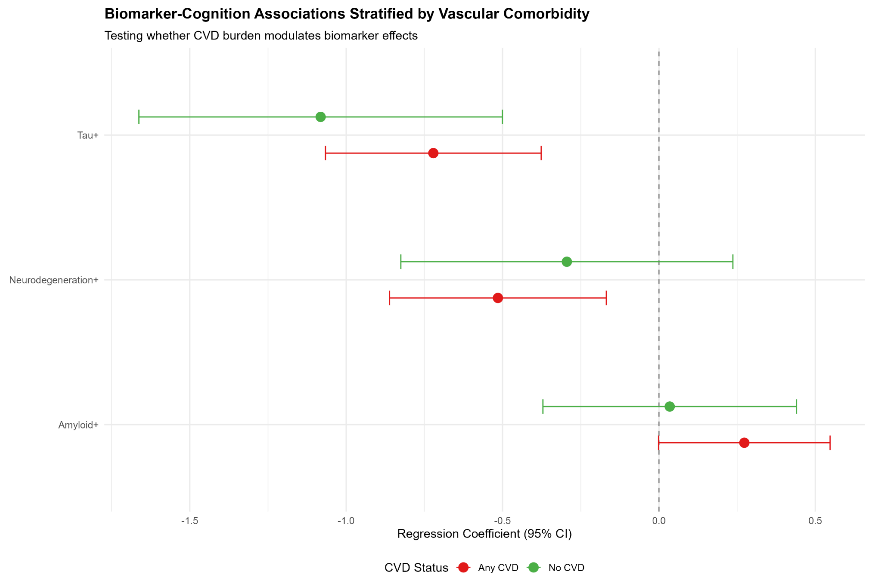


**Note:** Forest plot showing regression coefficients and 95% confidence intervals for biomarker-cognition associations stratified by CVD status. Two groups per biomarker (amyloid, tau, neurodegeneration): No CVD (n=1,341) in green, Any CVD (n=2,955) in red. Vertical dashed line at $\beta=0$ indicating null effect. **X-axis:** Regression Coefficient (95% CI). **Y-axis:** Biomarker type.

**Figure S2 Legend:** Forest plot displaying biomarker-cognition associations stratified by cardiovascular disease (CVD) presence. No CVD (green) includes participants without hypertension, diabetes, or stroke. Any CVD (red) includes participants with one or more vascular conditions. All models adjust for age, sex, race/ethnicity, and education. Outcome is cognitive score (TICSm). Tau demonstrates robust associations in both strata, with particularly strong effects in CVD-free individuals ($\beta=-1.08$, $p<0.001$), suggesting powerful AD-specific associations when competing vascular pathology is absent. Neurodegeneration shows significant associations only in the Any CVD stratum ($\beta=-0.52$, $p=0.004$), consistent with NfL/GFAP capturing both AD and vascular brain injury. Amyloid remains non-significant in both strata. These findings indicate that biomarker-cognition relationships persist independent of vascular burden, though effect magnitudes vary, supporting partially distinct AD and vascular pathways to cognitive decline.

#### Figure S3. Biomarker Calibration by Race/Ethnicity

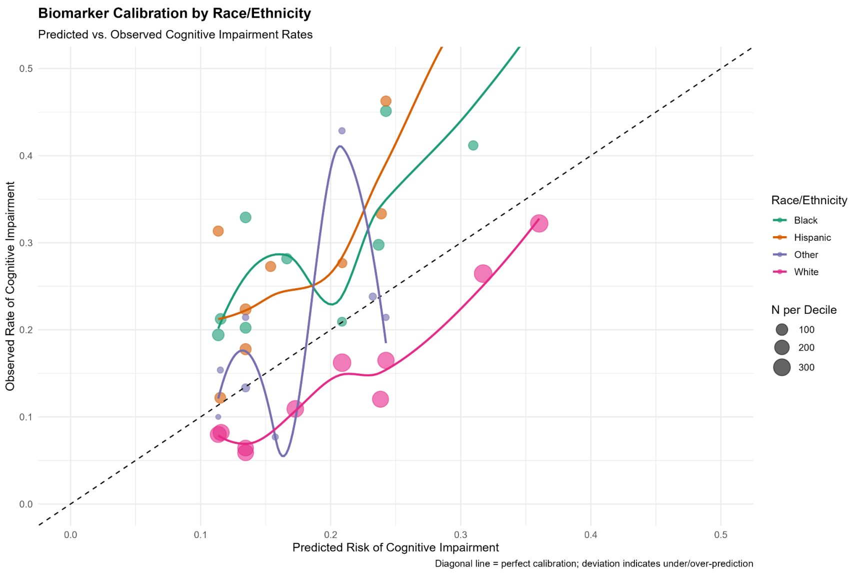


**Note:** Calibration plot with predicted risk of cognitive impairment (X-axis, 0-0.5) vs. observed rate of cognitive impairment (Y-axis, 0-0.5). Points represent risk deciles within each racial/ethnic group, sized by sample size, colored by race/ethnicity (White=pink, Black=teal, Hispanic=orange, Other=purple). Smooth loess curves fitted for each group. Diagonal dashed line indicating perfect calibration (predicted = observed). Points on the diagonal indicate good calibration; points above the diagonal indicate underprediction; points below the diagonal indicate overprediction.

**Figure S3 Legend:** Calibration plot comparing predicted vs. observed cognitive impairment rates across risk deciles, stratified by race/ethnicity. Predicted risk derived from logistic regression, including A+T+N biomarkers as predictors. Outcome is cognitive impairment (CIND or dementia). Each point represents a risk decile within a racial/ethnic group, with point size proportional to sample size (N per decile). Perfect calibration corresponds to points lying on the 45-degree diagonal (dashed black line); deviation indicates systematic under- or over-prediction. White participants (pink) show reasonable calibration with points clustering near the diagonal. Black (teal) and Hispanic (orange) participants show systematic miscalibration: observed rates exceed predicted rates at low risk (underprediction) and deviate unpredictably at high risk. Loess smoothing curves illustrate group-specific calibration patterns. These calibration failures indicate that biomarker-based risk models developed in predominantly White samples do not transport accurately to minoritized populations, necessitating race-specific recalibration or alternative modeling approaches to achieve equitable performance.

#### Figure S4. Race-Specific vs. Universal Optimal Biomarker Cutpoints

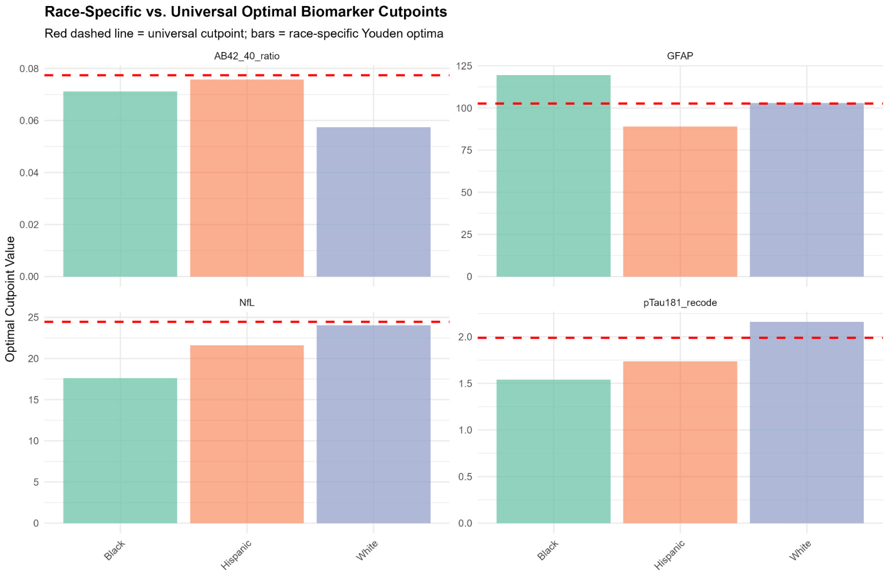


**Note:** Grouped bar chart with biomarkers (Aβ42/40, pTau181, NfL, GFAP) on X-axis, optimal cutpoint value on Y-axis. Bars colored by race/ethnicity (Black=teal, Hispanic=orange, White=purple). Horizontal red dashed lines indicate the universal (overall) optimal cutpoint for each biomarker. Faceted by biomarker with free Y-axis scales to accommodate different measurement units.

**Figure S4 Legend:** Comparison of race-specific Youden-optimized biomarker cutpoints vs. universal (overall population) thresholds. Bars represent optimal cutpoints for each racial/ethnic group derived by maximizing the Youden index (Sensitivity + Specificity – 1) within the group. Red dashed horizontal lines indicate universal cutpoints optimized across the entire sample. Substantial differences emerge: pTau181 optimal cutpoint varies from 1.54 pg/mL (Black) to 2.16 pg/mL (White), a 40% relative difference. NfL ranges from 17.6 pg/mL (Black) to 24.0 pg/mL (White), a 36% difference. GFAP shows a reversed pattern, with Black participants having a higher optimal threshold (119.5 pg/mL) than Hispanic (89.0 pg/mL) or White (102.9 pg/mL) participants. These empirically-derived race-specific optima suggest that universal cutpoints may systematically misclassify minoritized individuals, contributing to observed fairness disparities (12-percentage-point TPR gap). However, implementing race-specific thresholds raises ethical concerns about reifying biological race concepts and requires careful consideration of potential benefits vs. harms.

#### Figure S5. Fairness Landscape: TPR Across Race × Sex × Education

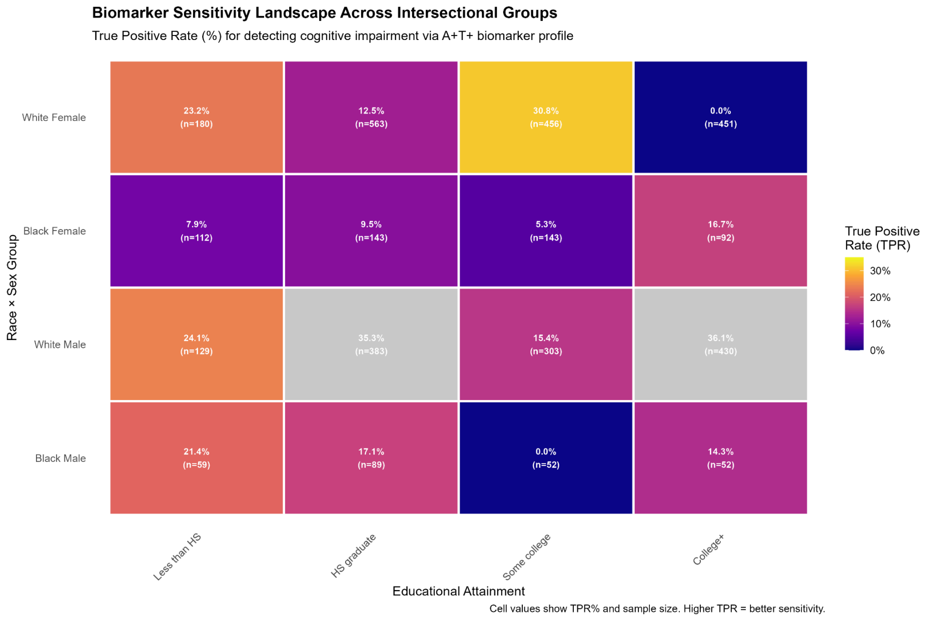


**Note:** Heatmap with Education level (Less than HS, HS graduate, Some college, College+) on X-axis and Race × Sex Group (White Female, Black Female, White Male, Black Male) on Y-axis. Cells colored by True Positive Rate (TPR) using plasma color scale (dark blue=0%, bright yellow=35%). Cell values display "TPR% (n=sample size)". Higher TPR indicates better biomarker sensitivity for detecting cognitive impairment.

**Figure S5 Legend:** Heatmap visualizing biomarker sensitivity landscape across intersectional demographic groups defined by race, sex, and educational attainment. True Positive Rate (TPR) represents the proportion of individuals with cognitive impairment correctly identified by the A+T+ biomarker profile. Color intensity indicates TPR magnitude (dark blue=low sensitivity, bright yellow=high sensitivity). Cell values show TPR percentage and sample size. Black women exhibit persistently low TPR across all education levels (range 0.0-16.7%), with particularly striking deficits in higher education strata (some college: 5.3%, college+: 16.7%). White men show the highest TPR across education levels (range 15.4-36.1%). These patterns demonstrate that educational attainment does not fully mitigate race/sex-based biomarker performance gaps, consistent with intersectionality theory positing that multiple marginalized identities produce unique forms of disadvantage not reducible to additive effects. The finding that Black women with a college education show TPR=16.7% (vs. White men with less than HS showing TPR=24.1%) illustrates how racism and sexism compound to produce systematic biomarker performance disparities even among highly educated minoritized individuals.

#### Figure S6. Conceptual Framework: Equity and Transportability in Plasma ATN Biomarkers

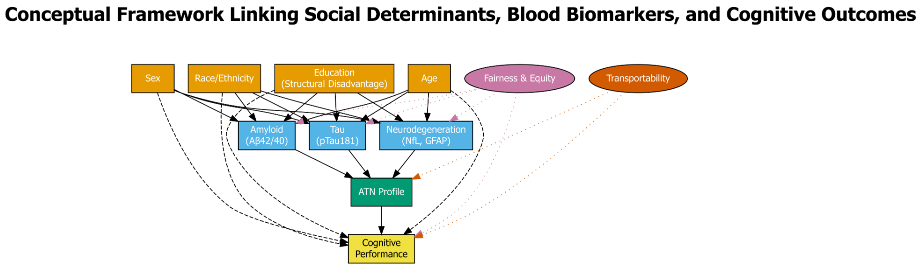


**Figure S6 Legend:** Directed acyclic graph (DAG) illustrating the conceptual model guiding equity-centered analyses. Yellow boxes represent demographic factors and structural determinants (race/ethnicity, sex, education as proxy for structural disadvantage, age). Blue boxes represent plasma biomarkers (amyloid [Aβ42/40], tau [pTau181], neurodegeneration [NfL, GFAP]) and derived ATN profile classification (green box). The yellow box at the bottom represents the primary outcome (cognitive performance). Solid arrows indicate direct causal pathways or associations; dashed arrows indicate modification effects (demographics modify biomarker-cognition relationships). Pink and orange ellipses represent analytic frameworks (Fairness & Equity, Transportability) applied to evaluate biomarker performance and generalizability. Dotted arrows from frameworks to biomarkers and cognition indicate these lenses examine equity and transportability across all pathways. This framework emphasizes that biomarker interpretation cannot be divorced from social context; structural disadvantage operates "under the skin" to modify dementia pathophysiology.

**Figure S7. Sensitivity Analysis: ATN Prevalence Across Biomarker Cutpoint Definitions**

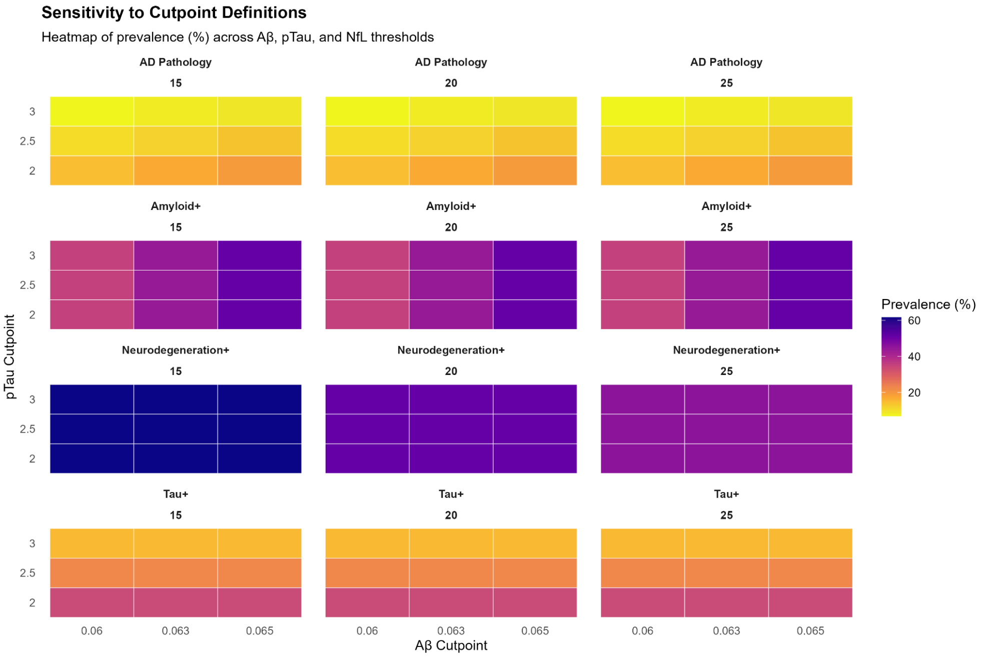


**Figure S7 Legend:** Heatmap grid displaying prevalence (%) of amyloid positivity (A+), tau positivity (T+), neurodegeneration positivity (N+), and AD pathology (A+T+) across 27 combinations of biomarker cutpoints. Rows represent pTau181 thresholds (2.0, 2.5, 3.0 pg/mL), columns represent Aβ42/40 thresholds (0.060, 0.063, 0.065), and panels represent NfL thresholds (15, 20, 25 pg/mL). Color intensity indicates prevalence magnitude (yellow=low, purple=high). AD pathology prevalence varies 2.8-fold from 6.8% (most stringent: Aβ <0.060, pTau >3.0, NfL >25) to 19.2% (most lenient: Aβ <0.065, pTau >2.0, NfL >15). This dramatic sensitivity underscores the critical need for consensus thresholds validated against neuropathology in diverse populations. Small changes in cutpoints produce large shifts in prevalence with implications for screening programs, clinical trial eligibility, and public health burden estimates. The intermediate combination (Aβ <0.063, pTau >2.5, NfL >20) used in the main analyses yields 12.1% AD pathology, closely matching the literature consensus.

### Figure S8. Biomarker Distributions by Education Level

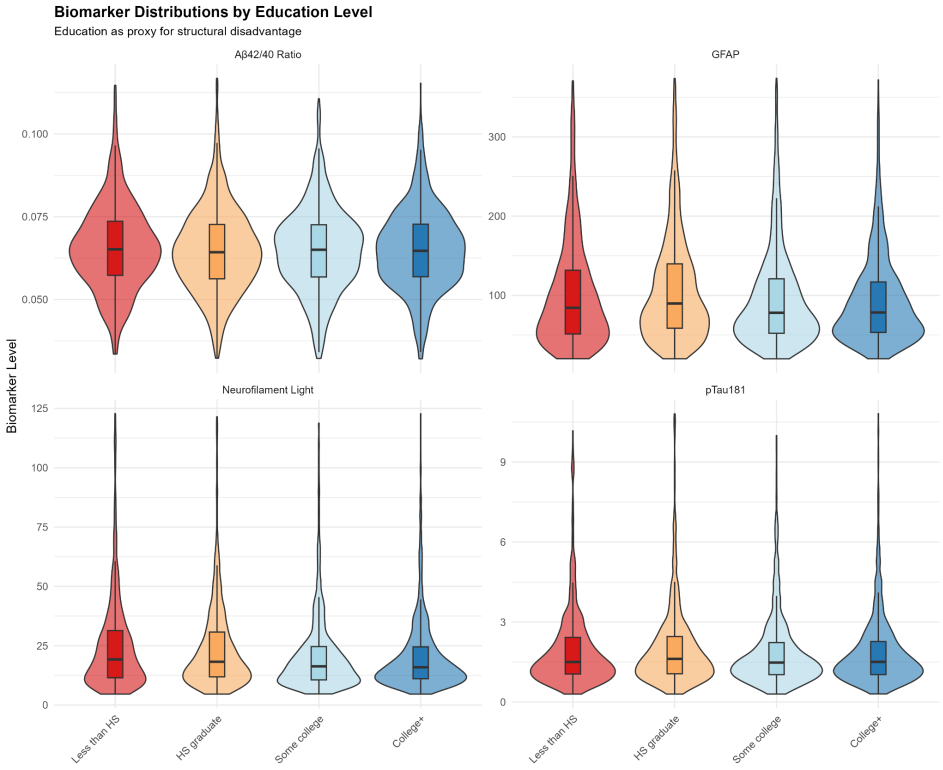


**Figure S8 Legend:** Violin plots with overlaid boxplots showing distributions of four plasma biomarkers (Aβ42/40 ratio, GFAP [pg/mL], neurofilament light [NfL, pg/mL], pTau181 [pg/mL]) across education levels (Less than HS, HS graduate, Some college, College+). Data trimmed to 1st-99th percentiles to reduce outlier influence on visualization. Inverse gradients emerge, with lower education being associated with lower Aβ42/40 ratios (indicating more amyloid pathology) and higher NfL/GFAP levels (suggesting more neurodegeneration), supporting the hypothesis that structural disadvantage confers a neuropathological burden. These distributions align with the "brain maintenance" hypothesis: higher education confers neuroprotection through cognitive reserve, vascular health, and reduced inflammation. Median values shift systematically across education strata, although substantial overlap exists, indicating that education serves as a proxy for complex life-course socioeconomic exposures rather than a deterministic biological variable. Educational gradients in biomarkers illuminate pathways through which social inequality becomes embodied as neurobiological differences.

#### Figure S9. Biomarker-Cognition Associations by Education Level (Forest Plot)

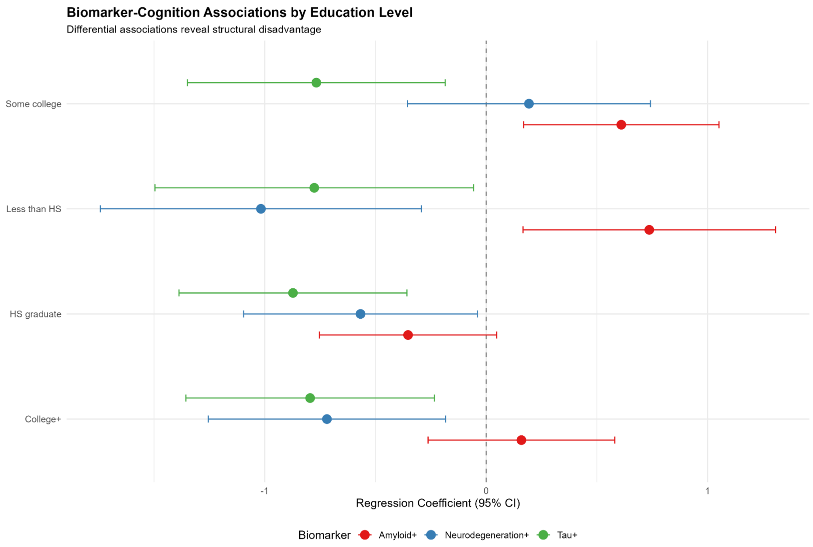


**Figure S9 Legend:** Forest plot displaying regression coefficients (β) and 95% confidence intervals for biomarker-cognition associations, stratified by education level. Each panel represents an education stratum (Less than HS, HS graduate, Some college, College+), with separate estimates for amyloid (red), neurodegeneration (blue), and tau (green). Dashed vertical line at β=0 indicates null effect. Paradoxical positive amyloid associations emerge in low-education groups, while neurodegeneration shows the strongest negative effects in structurally disadvantaged populations. Tau demonstrates consistent negative associations across all education levels.

#### Figure S10. Biomarker Distributions by Race × Sex Intersectional Groups

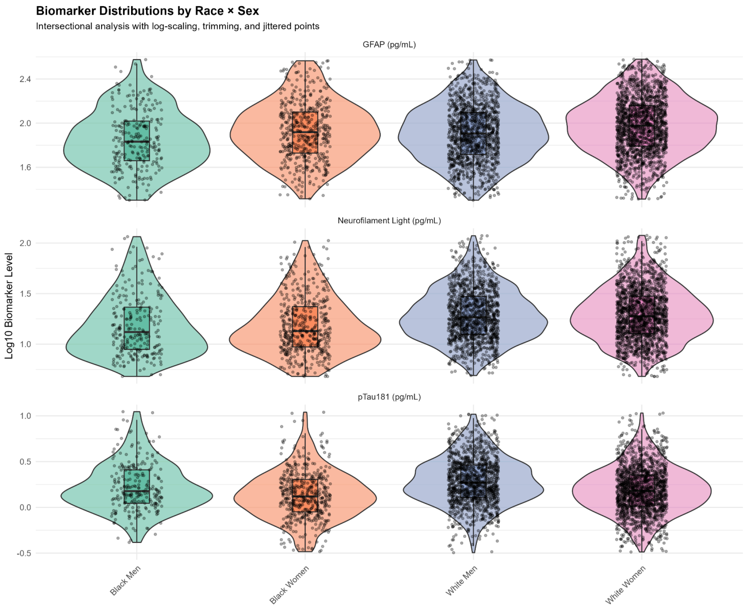

**Figure S10 Legend:** Violin plots with overlaid boxplots and jittered individual data points (log10-transformed) for three plasma biomarkers (GFAP, neurofilament light [NfL], pTau181) across four intersectional demographic groups (Black Men, Black Women, White Men, White Women). Substantial within-group variability and between-group overlap characterize all biomarkers. Subtle elevation in NfL is observed in Black men relative to other groups. Data are trimmed to 1st-99th percentiles and log-transformed for visualization. This distribution overlap complicates the establishment of universal biomarker thresholds applicable across diverse populations.

#### Figure S11. Biomarker Performance (AUC) by Race × Sex

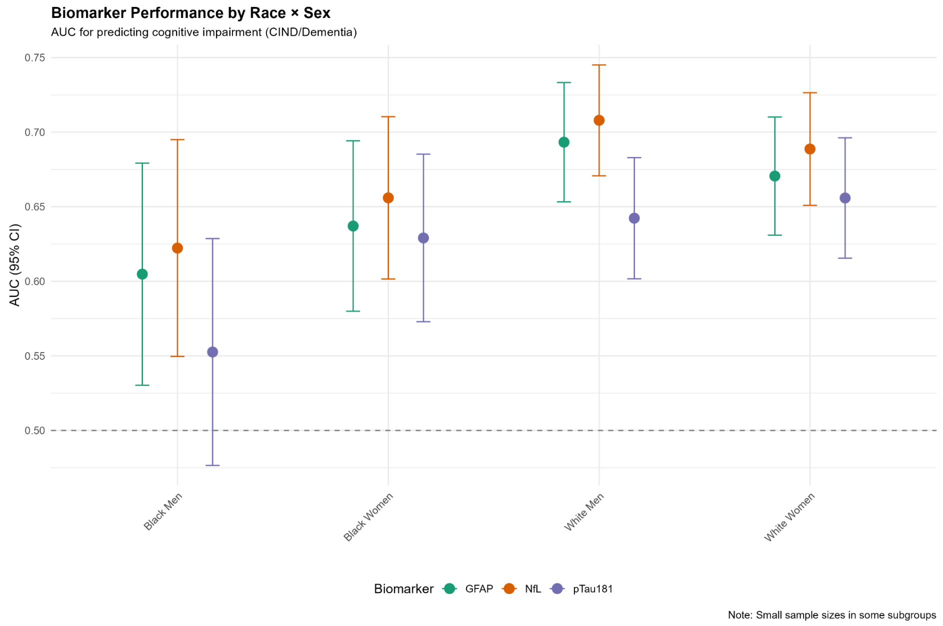


**Figure S11 Legend:** Forest plot displaying area under the receiver operating characteristic curve (AUC) with 95% confidence intervals for three biomarkers (GFAP [turquoise], NfL [orange], pTau181 [purple]) predicting cognitive impairment (CIND or dementia), stratified by race × sex intersectional groups. Dashed horizontal line at AUC=0.50 indicates chance performance. White participants demonstrate superior biomarker discrimination (AUC 0.64-0.71) compared to Black participants (AUC 0.55-0.66), with Black men's pTau181 AUC (0.55) barely exceeding chance. Small sample sizes in some subgroups yield wide confidence intervals. Note indicates caution in interpretation due to sample size limitations.

#### Figure S12. Biomarker Missingness Patterns by Demographics

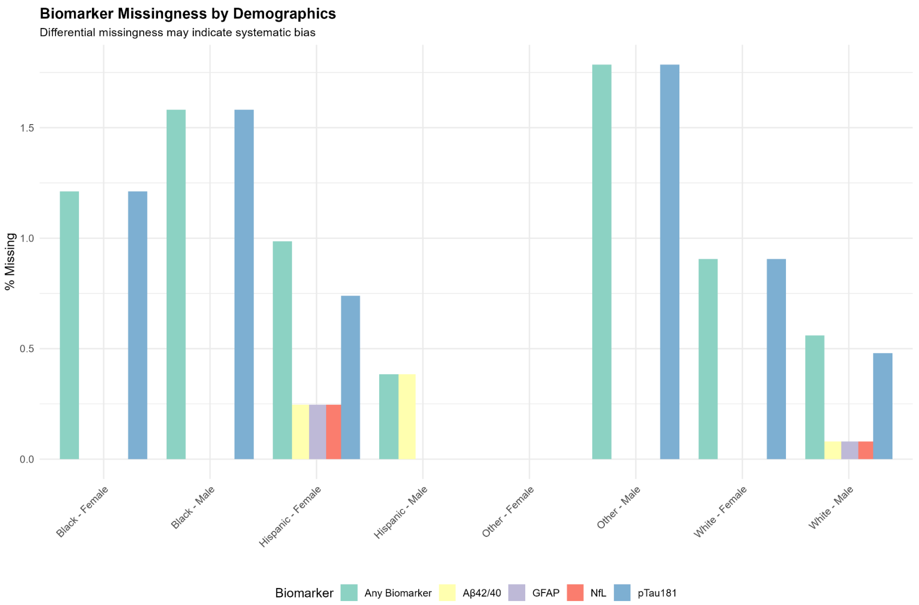


**Figure S12 Legend:** Grouped bar chart displaying percentage of missing biomarker data (any biomarker, Aβ42/40, GFAP, NfL, pTau181) across race × sex demographic groups. Bars are color-coded by biomarker type. Differential missingness is evident, with Hispanic participants showing the highest Aβ42/40 missingness and pTau181 showing the most frequent missingness across groups. Black and White participants show relatively low missingness (<2%), while "Other" groups show virtually complete data. These patterns may reflect systematic differences in sample handling, assay performance, or participant characteristics.

#### Figure S13. Race × Amyloid Interaction on Cognition

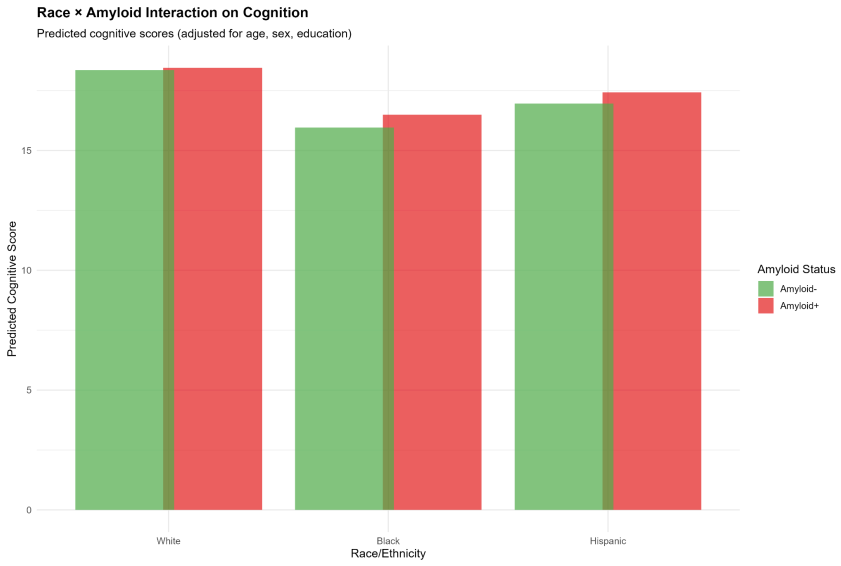

**Figure S13 Legend:** Grouped bar chart displaying predicted cognitive scores (TICSm) by race/ethnicity (White, Black, Hispanic) and amyloid status (Amyloid- [green], Amyloid+ [red]), adjusted for age, sex, and education. Among White participants, amyloid positivity shows minimal cognitive impact (difference <0.2 points). Among Black and Hispanic participants, amyloid positivity is associated with lower predicted cognition, though confidence intervals overlap. These differential associations suggest that biomarker-cognition relationships are not uniform across racial/ethnic groups, potentially reflecting biological heterogeneity or measurement bias.

#### Figure S14. Race × Neurodegeneration Interaction on Cognition

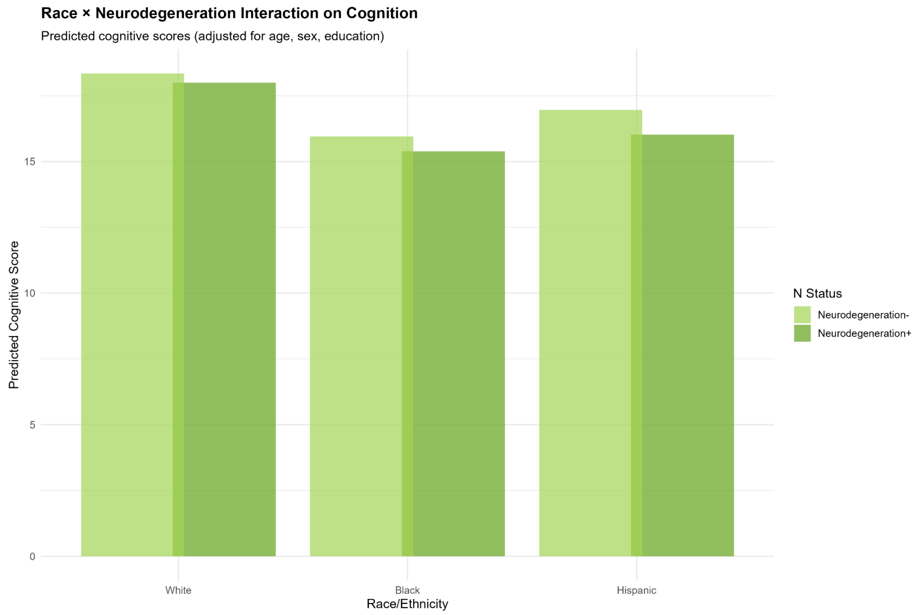


**Figure S14 Legend:** Grouped bar chart displaying predicted cognitive scores (TICSm) by race/ethnicity (White, Black, Hispanic) and neurodegeneration status (N- [light green], N+ [dark green]), adjusted for age, sex, and education. Neurodegeneration positivity is consistently associated with lower cognitive scores across all racial/ethnic groups, though the effect magnitude varies. White and Hispanic participants show similar patterns, while Black participants demonstrate somewhat attenuated associations. These relatively consistent effects suggest neurodegeneration markers (NfL, GFAP) may transport more reliably across populations than amyloid.

#### Figure S15. Race × Tau Interaction on Cognition

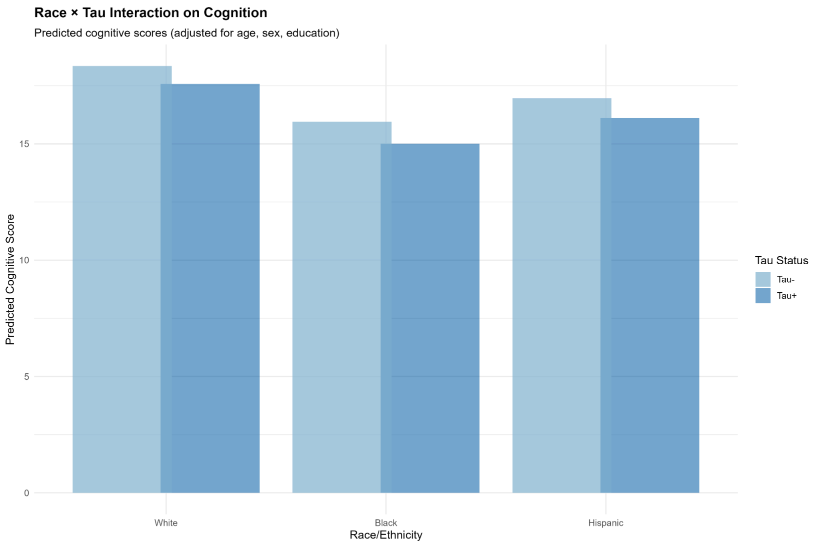


**Figure S15 Legend:** Grouped bar chart displaying predicted cognitive scores (TICSm) by race/ethnicity (White, Black, Hispanic) and tau status (Tau**-** [light blue], Tau+ [dark blue]), adjusted for age, sex, and education. Tau positivity is associated with lower cognition across all racial/ethnic groups with similar effect magnitudes (approximately 1-point reduction). This consistency supports tau as the most transportable plasma biomarker for cognitive prediction at the population level, aligning with neuropathological literature demonstrating strong correlations between neurofibrillary tangles and clinical dementia severity.
